# Learning good therapeutic targets in ALS, neurodegeneration, using observational studies

**DOI:** 10.1101/2024.10.11.24315263

**Authors:** Mohammadali Alidoost, Jeremy Y. Huang, Georgia Dermentzaki, Anna S. Blazier, Giorgio Gaglia, Timothy R. Hammond, Francesca Frau, Mary Clare Mccorry, Dimitry Ofengeim, Jennifer L. Wilson

**Author notes:** To whom correspondence should be addressed: Jennifer L Wilson, 420 Westwood Plaza, 5121 Eng V, Rm 4121D, Los Angeles CA, 90095, USA.

## Abstract

Analysis of real-world data (RWD) is attractive for its applicability to real-world scenarios but RWD is typically used for drug repurposing and not therapeutic target discovery. Repurposing studies have identified few effective options in neuroinflammatory diseases with relatively few patients such as amyotrophic lateral sclerosis (ALS), which is characterized by progressive muscle weakness and death with no disease-modifying treatments available. We previously reclassified drugs by their simulated effects on proteins downstream of drug targets and observed class-level effects in the EHR, implicating the downstream protein as the source of the effect. Here, we developed a novel ALS-focused pathways model using data from patient samples, the public domain, and consortia. With this model, we simulated drug effects on ALS and measured class effects on overall survival in retrospective EHR studies. We observed an increased but non-significant risk of death for patients taking drugs associated with the complement system downstream of their targets and experimentally validated drug effects on complement activation. We repeated this for six protein classes, three of which, including multiple chemokine receptors, were associated with a significant increased risk for death, suggesting that targeting proteins such as chemokine receptors could be advantageous for these patients. We recovered effects for drugs associated with complement activation and chemokine receptors in Parkinson’s and Myasthenia Gravis patients. We demonstrated the utility of network medicine for testing novel therapeutic effects using RWD and believe this approach may accelerate target discovery in neuroinflammatory diseases, addressing the critical need for new therapeutic options.

## Introduction

We need novel treatment identification methods for rare, difficult-to-treat diseases, especially those with insufficient experimental systems. Big data approaches and advanced algorithms have the power to capture patient data and infer new disease drivers in ways previously not possible. Many approaches suffer from limitations that were well-established a decade ago: animal models are valuable for understanding mechanism but have limited relevance to human conditions, human epidemiology is highly relevant to human disease but is insufficient for explaining causality, and natural conditions are the most relevant and may explain causality but are rare^1^. We hypothesized that a model-informed analysis of patient data may overcome these hurdles, by predicting causality first, and later testing causal predictions in relevant human disease contexts, thereby overcoming the limitations of traditional approaches.

Retrospective analyses of electronic health records (EHRs) are attractive for difficult-to-treat conditions, including many neuroinflammatory conditions, where animal and model systems have failed to deliver treatments with substantial impacts on survival. However, when inferring treatment impact from real-world evidence, most studies are restricted to repurposing of known druggable targets. For instance, the discovery of phosphodiesterase inhibitors as protective for Parkinson’s^2^ and amyotrophic lateral sclerosis (ALS)^3^, peroxisome proliferator-activated receptor (PPAR) agonists as protective in Alzheimer’s Disease^4^, and the potential of multiple drugs, diazoxide, gefitinib, paliperidone, and dimethyltryptamine as high-quality candidates for repurposing in ALS^5^ all leverage ‘omics data, network medicine, and EHRs, but they uncovered evidence for known, druggable targets. Indeed, reviews of the field emphasized that real world data can support clinical trial emulation, sub phenotyping, or image analysis^6^, but the authors failed to consider the historical record as sufficient for supporting *novel, untested* drug targets. Despite the status quo, we had compelling prior evidence that it might be possible to more directly vet novel targets in EHRs and sought to test that idea in difficult-to-treat neuroinflammatory populations. Specifically, we learned that disease-specific network models improved prediction of drug effects^7^ and that drug-effect predictions based on shared protein networks were detectable in EHRs^8^.

Debilitating neuroinflammatory conditions without substantially effective treatments have motivated creative and compelling approaches to advance treatment options. Of the seven approved treatments for ALS by the Federal Drug Administration (FDA), none are disease-modifying^9^ and none prevent death. Multiple studies have identified distinct molecular mechanisms including the role of the innate immune system (**Table S4**), among other genetic drivers ^9,10^. Targeting these disrupted pathways or causal genetic drivers in the clinic has had limited efficacy and coverage due to the time to develop new medicines and initiate new trials. Since ALS is a rare disease, clinical trial cohorts often struggle to meet statistically significant thresholds, leading to increased interest in leveraging new insights from real-world-data. Thus, we pursued our approach in neuroinflammatory conditions because of the potential for accelerating discovery for these devastating conditions.

In this study, we tested our network inference platform on neuroinflammatory conditions and corroborated our model predictions EHR investigation with experimental analysis. Intriguingly, our results synergize with other investigations of dysregulated pathways in ALS, specifically, we recovered increased risks for death associated with complement system activation. And despite the fact that ALS patients were not prescribed any complement-targeted drugs, our retrospective analysis provided rationale for directly inhibiting this pathway. We also asked whether these insights were generalizable across additional neuromuscular and neurodegenerative diseases with larger patient populations, such as Myasthenia gravis and Parkinson’s disease. In total, we built and tested six network classes with different observed significant impacts on overall patient survival in several modules. Encouragingly, our pipeline independently identified therapeutic targets currently in clinical development and suggested additional targets with stronger effects. By classifying approved therapies by their effects on downstream proteins, rather than labeled primary targets, we can leverage large-scale cohort studies to identify novel targets for ALS and other neurodegenerative diseases.

## Materials and Methods

### Curating ALS-associated neurogenerative disease pathways

To customize our network analysis, we curated novel disease pathways from multiple sources. All together, we included 195,252 gene-phenotype relationships across 1,096 unique disease pathways and 24,441 unique genes/transcripts. We curated this data from six sources listed below. Further, a list of all derived pathway names, their sources, and the total number of genes is included in **Supplemental File 2**. For a detailed list of pathway sources and processing for each source, and integration into our network modeling platform please see *Supplemental Methods*.

### Running ALS-PathFX on all approved drugs

Like previous versions of PathFX, the ALS version required drug binding proteins as inputs. We used drug target information from DrugBank version 5.1.6. as inputs for ALS-PathFX analysis. We generated networks and association tables for all drugs and then searched the PathFX-generated association tables for phenotypes of interest. The association tables are standard outputs from PathFX and included all network-associated phenotypes, the multiple-hypothesis-corrected p-values for the associations, and which disease pathway proteins were discovered in the networks. All ALS-PathFX predictions, created by merging all associations tables for drugs analyzed, are included in **Supplemental File 2**.

### Generating network classes

Using the merged table of all ALS-PathFX drug-phenotype associations, we counted how often a network protein was used to make a drug-phenotype prediction (e.g., CXCL13 was used to predict a network association from gabapentin to the “MS CSF” and “Bulk RNAseq ALS vs Control” pathways, and C3 was used to predict a network association from gabapentin to the “Bulk RNAseq ALS vs Control” pathway). After identifying network proteins occurring in the most drug-ALS PathFX associations, we developed new drug classes by grouping drugs that shared network proteins (e.g., CXCL13 or C3). We completed this analysis using the pandas module in python. Because we wanted to study patient data, we also filtered out experimental or unapproved drugs and counted the number of approved active ingredients with each network protein.

In a few instances, we observed several related network proteins and made multi-protein classes based on qualitative and quantitative assessment of the ranked proteins, such as in the case for the complement system proteins and chemokine proteins. Grouping drugs based on network proteins yielded groups for the “target” cohorts (“network” class). To generate comparable “comparator” cohorts, we assessed all remaining approved drugs that had network associations to the same ALS phenotypes, but lacked a network protein in the association (“non-network” class). For instance, all drugs with an association to the “MS CSF” or “Bulk RNAseq ALS vs Control” pathways would be considered in the non-CXCL13 comparator class. All network proteins and their presence in drug networks are contained in **Supplemental File 3**). Code for this analysis is included in our GitHub.

### Clinical data – Optum Market Clarity

We used the Optum’s Market Clarity data: Optum® Market Clarity Data is an innovative dataset of integrated multi-source medical and pharmacy claims and electronic health record (EHR) data that enables detailed evaluation and assessment of the patient journey. The Market Clarity Data Asset combines robust transactional pharmacy and medical claims data with best-of-breed EHR data. It is a fully HIPAA-compliant, statistician-certified, de-identified precision data set that uncovers hidden intelligence to turn insights into quick action. The Market Clarity Dataset links EHR data with historical, linked administrative claim data, pharmacy claims, physician claims, facility claims (with clinical information) and is inclusive of medications prescribed and administered. Clinically rich and specific data elements sourced from the EHR include lab results, vital signs and measurements, diagnoses, procedures and information derived from unstructured clinical notes using natural language processing.

We specifically assessed patient records available through February 2023 and accessed the following data tables – the membership info for collecting date of birth, sex, race, ethnicity, and gender, when available; the RX claims table to assessing filled prescriptions as a proxy for drug exposures; the diagnosis table for identifying ALS patients and other comorbid conditions; and the date of death table for collecting the month/year of death, when available.

### Observational studies in the electronic health record

To measure effects for our network-derived drug classes, we used a standard target-comparator approach using best practices in propensity score estimation and weighted treatment effects. We’ve used a similar approach in our own work^8^ and describe the approach in greater detail in *Supplemental Methods*.

### Stem Cell Culture

Human iPSC astrocytes (iCell astrocytes, Fujifilm, Cat# C1037) were cultured in Matrigel-coated (Corning 354277) 96-well plates with DMEM/F12 (Life Technologies 11330057) media supplemented with heat-inactivated FBS (Gibco A38400-01) and N-2 supplement (Gibco 17502-048).

### Stimulation and drug treatment

Three days post-plating, iCell astrocytes were treated with selective compounds (**Table S2**) at 100 nM, 1µM, and 10 µM concentrations. For astrocyte stimulation (A1 subtype) we used the A1 cocktail: IL-1a (R&D Systems, Cat# 200-LA, 0.3 ng/mL), TNF alpha (R&D Systems, Cat# 210-TA, 3 ng/mL) and C1q (MyBiosource, Cat# MBS147305, 40 ng/mL) factors. Both the compounds, at multiple doses (100 nm, 1 uM, and 10 uM), and the A1 cocktail factors were added simultaneously on the cultures for overnight treatment (24h).

### ELISA for complement system activation

Media were collected 24h post-treatment and complement levels were assessed with the Milliplex Human Complement Panel 1 (Millipore, HCMP1MAG-19K / Factors: C1q, C3, C3b/iC3b, C4, Factor B, Factor H) and 2 (Millipore, HCMP2MAG-19K / Factors: C2, C4b, C5, C5a, Factor D, Mannose-binding lectin (MBL), Factor I) according to the kit’s instructions. The assay was run in a Bio-Plex 2000 instrument (Biorad).

### Data availability

The Market Clarity data used were licensed from Optum and are not publicly available. To increase the rigor and reproducibility of our work, we have released code and data for the network analysis, and anonymized code for accessing clinical data through a GitHub repository: https://github.com/jenwilson521/network_drug_classes_als/ All supplemental Excel files are included in their original format in the GitHub repository to increase readability and reusability of the data.

## Results

### ALS-PathFX predicted drug network associations to ALS through distinct proteins

Our overall approach used pathway modeling to identify novel drug classes and observational studies in the health record to test the utility of these drug classes (**Figure 1**). To build a custom network medicine platform, we curated several high-quality datasets from the TargetALS data portal and several patient-derived datasets^11^. The TargetALS datasets included differential genes measured by RNAseq from multiple tissue types including cerebellum, motor cortex, and spine. We derived multiple gene sets from this data using different fold-change thresholds (see *methods*). We also leveraged multiple novel sequencing and proteomics data derived from patient samples, including snRNAseq in multiple tissue types, proteomics from patient cerebral spinal fluid (CSF), and RNAseq data from reactive astrocytes and microglia (for a full list see *methods* and^11^). We specifically modified our highly-flexible PathFX platform (released with ^12^) to emphasize these gene lists (referred to as “pathway phenotypes”) into PathFX^12–14^ using the original bias reduction methods published in ^13^ and described in methods. We then used our custom network algorithm, ALS-PathFX, to model drug effects and derive new drug classes based on predicted effects on nearby, but untargeted (not bound by drug) proteins (**Figure 1B**). Taken together, our ALS-PathFX network model leverages several high-quality disease-relevant datasets to make predictions about drug effects.

**Figure 1.**
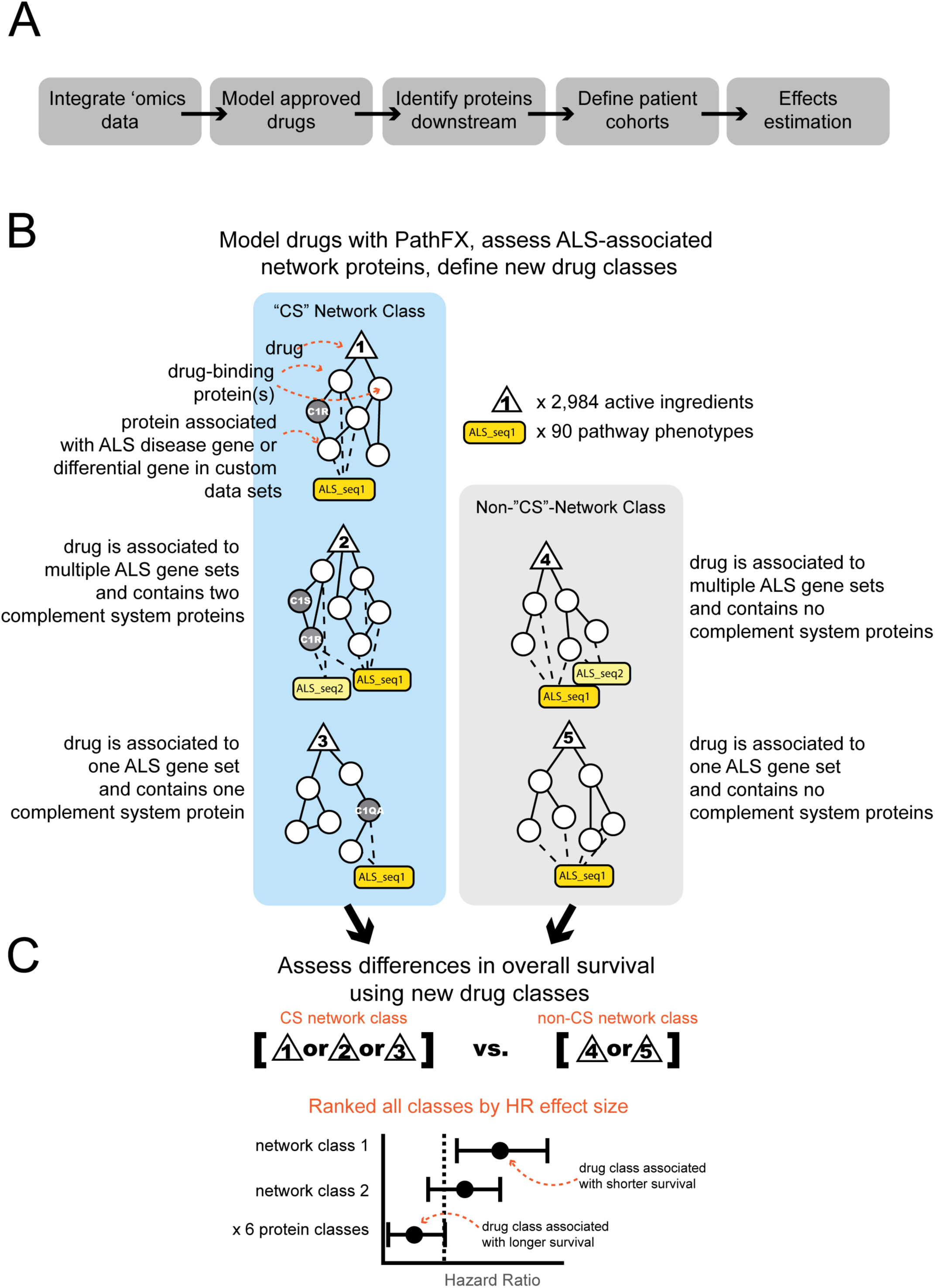
A combined pathways and observational study approach for identifying pathways with protective or harmful effects on overall survival in ALS. (**A**) Our overall workflow consisted of ‘omics data integration using several high-quality data sets, network modeling, identification of downstream proteins, identifying patient cohorts, and effects estimation (**B**) For network inference, we modeled all approved, active ingredients using ALS-PathFX and then created novel drug classes based on downstream proteins included in drug networks. In this example, we assessed drugs with network associations to multiple ALS sequencing datasets and included them in the “complement system” (“CS”) drug class if they had any complement-system proteins in their networks. We highlighted five drugs that are associated with any one of two disease pathways (i.e., “ALS sequencing data 1”, or “ALS sequencing data 2”). In our schematic, we also highlighted that drugs “1”, “2”, and “3” contain downstream associations to complement-system proteins and drugs “4” and “5” are connected to the same pathway phenotypes, but through different proteins. Drugs, network proteins, and disease phenotypes are represented by triangles, circles, or yellow boxes, respectively. (**C**) Later, we measured overall survival for patients exposed to at least one of the “complement class” or “non-complement class” drugs using observational studies in the Optum Market Clarity dataset. In this example, we grouped drugs “1”, “2”, and “3” into the “target” class, and drugs “4” and “5” into the “comparator” class. We repeated this analysis for 6 drug classes and ranked classes by their effects on overall survival. Three complement proteins are shown in the diagram, but a total of nine were considered when assigning drugs to the complement system class: “Complement component 1, R subcomponent (C1R)”; “Complement component 1, S subcomponent (C1S)”; “Complement component 1, Q subcomponent, alpha polypeptide (C1QA)”; “Complement component 1, Q subcomponent, beta polypeptide (C1QB)” “Complement component 1, Q subcomponent, alpha polypeptide (C1QA)”; “Complement component 1, Q subcomponent, gamma polypeptide (C1QC)”; “Complement C3 (C3)”; “Complement C5 (C5)”; “Complement C3a receptor 1 (C3AR1)”; & “Complement C5a receptor 1 (C5AR1)”.

We generated ALS-PathFX networks for 7,013 approved and experimental drugs using targets from DrugBank^15,16^. Briefly, ALS-PathFX first identified high-confidence proteins downstream of druggable targets, using interaction specificity analysis, a process that prioritizes protein interactions while reducing connections through hub proteins. After generating a protein network for each drug, ALS-PathFX identified network-associated phenotypes, by assessing which disease pathway proteins were enriched in the drug network relative to the entire interactome, and using multiple hypothesis correction and controlling for biases in the number of pathway-associated genes/proteins (as described in ^13^ and in *methods*). In total, we discovered 13,967 drug-phenotype associations containing 2,984 unique drugs and 90 unique disease pathways. For example, PathFX generated a network of 235 proteins and 238 edges for the antipsychotic, acetophenazine, and connected the network to 138 phenotypes, including 4 neurodegenerative phenotypes (subnetwork associated with four phenotypes shown in **Figure 2**).

**Figure 2.**
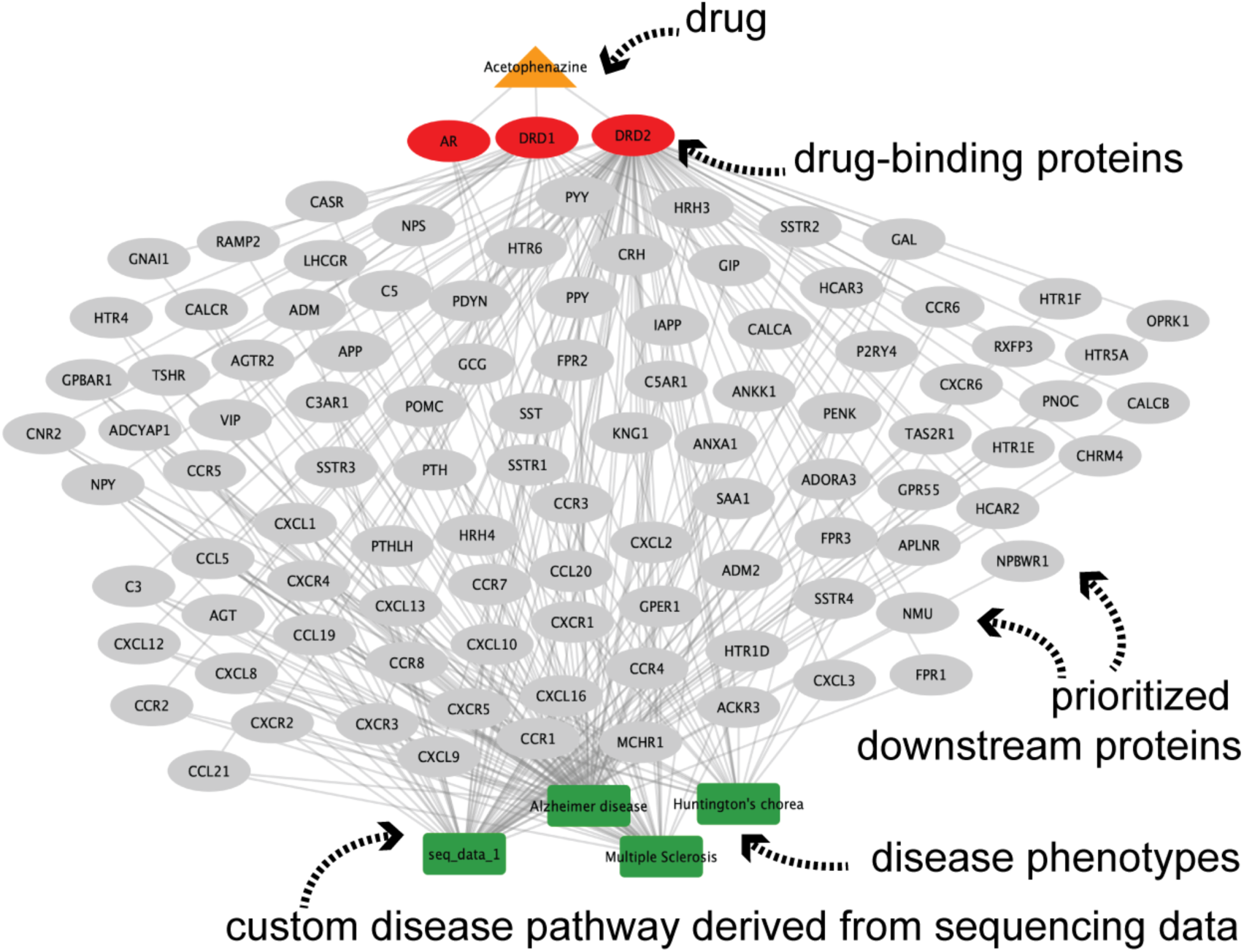
ALS-PathFX connected acetophenazine to multiple neurodegenerative phenotypes. PathFX connected the drug, acetophenazine (orange triangle) to four relevant neurodegenerative phenotypes (green boxes): Alzheimer disease, Huntington’s chorea, seq_data_1, and Multiple Sclerosis. PathFX used three drug target proteins (red circles): androgen receptor (AR), and the dopamine receptors D1 and D2 (DRD1, DRD2), and 99 adjacent proteins (gray ellipses) when predicting associations to these phenotypes.

Acetophenazine’s network contained the complement system proteins Complement C3 (C3), Complement C5 (C5), Complement C5a Receptor 1 (C5AR1), and Complement C3a Receptor 1 (C3AR1). Network proteins may be associated with more than one phenotype, and for instance, C3 and C3AR1 were associated with “Multiple sclerosis” and “sequencing data 1”. We assigned acetophenazine to the “complement-associated drug class” or “CS-class” for brevity. We considered any drugs predicted to affect the same phenotypes (e.g., “Multiple sclerosis” or “sequencing data 1”) but without complement proteins in their networks, as sufficient comparator drugs and considered them in the “non-CS-class” of drugs. We provide more detail about ALS-PathFX results and assessment of network-protein drug classes in the *Supplemental Results*.

### Complement-system-associated drugs are associated with changes in survival

We hypothesized that drugs with complement system proteins in their networks would have distinct clinical effects. This hypothesis is further supported by multiple recent developments of complement system inhibitors for neurodegenerative disease, specifically myasthenia gravis (**Table S4**). Aside from recent discoveries, complement system inhibitors have been approved since 2007 for several other non-neurodegenerative diseases including autoimmune and blood disorders (**Table S4**). Despite several approved compounds, none were prescribed in our ALS study patient population (*described below and depicted in* **Figure S6A**, all compounds listed in **Supplemental File 4**), further emphasizing the need for a way to anticipate their effects on overall survival.

To test whether ALS-PathFX-predicted drugs affected overall survival, we conducted a novel observational study a using standard treatment-comparator study design and large-scale propensity scoring to adjust for possible confounding variables. More specifically, we accessed the de-identified Optum Market Clarity data. Optum Market Clarity deterministically links medical and pharmacy claims with EHR data from providers across the continuum of care. This data is available for researchers without IRB approval (*longer description in Methods*). The novelty in our approach was deriving network-protein-based drug classes instead of using traditional drug classifications such as anatomical therapeutic category (ATC) classification systems. We provide a more intuitive description of this process in the *Supplemental Results*.

After controlling for possible confounding (**Figure S6**), we estimated the differences in overall survival and observed a marginal, decreased, but not significant survival time for patients with an exposure to CS-class drugs (HR = 1.031 95% confidence interval (0.937, 1.134), p = 0.534) (**Figure 3A**, **Table 1**). The study discovered a detrimental effect early, where CS-class drugs are separated from non-CS-class drugs before ∼2,000 days (∼5.5 years). At later timepoints, ∼10,000 days (∼27.4 years), overall survival was minimally distinguishable. While the result is modest and insufficient to directly change clinical practice, this result affirmed that network-derived drug classes could predict differential effects on ALS survival in historical patient cohorts and suggested the possibility of testing other network drug classes. Yet, as mentioned previously, the results do not confirm whether drugs activate or inhibit the complement system.

**Figure 3.** Patient cohorts in network drug classes have sufficient covariate balance. (**A**) We plotted the total exposure days (sum of all prescriptions) per brand name per patient. We repeated this process for the target, CS-class (left) and comparator, non-CS-class (right) drugs. Each dot is a single patient. (**B**) The number of patients is plotted against their propensity score for patients on non-CS-class drugs (left) or CS-class drugs (right). The propensity score is the predicted probability using LogisticRegression on patient demographic, diagnostics, and medical prescription claims features. Patient demographics information for race (**C**) and gender (**D**) concepts.

**Figure 3.**
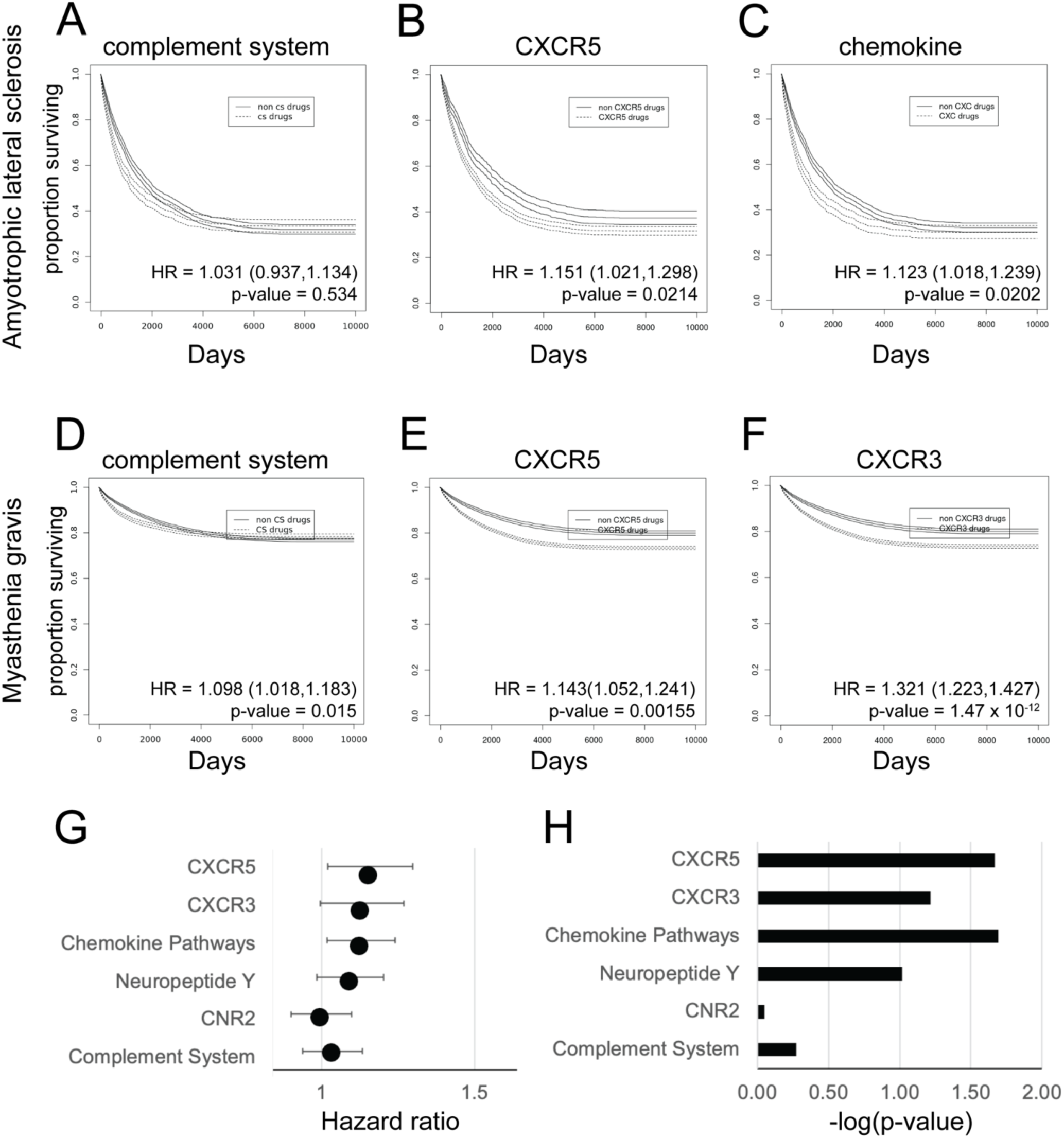
Multiple network drug classes have effects on overall survival in ALS, myasthenia gravis. We plotted Kaplan Meier survival curves for the complement system class (**A**), the chemokine receptor 5 (CXCR5) class, a relatively high-risk class (**B**), and the chemokine pathway class, also a relatively-high-risk class (**C**) in ALS patients. We also plotted Kaplan Meier plots for three network classes in myasthenia gravis patients: the complement system (**D**), chemokine receptor 5 (CXCR5) (**E**), and the chemokine receptor 3 (CXCR3) (**F**) classes. In all plots, the proportion of the surviving population plotted against time in days. Patients exposed to a “network class” or “Non-network class” drug are represented with a dotted or solid line, respectively. Hazard ratio values with the 95% confidence interval for six network-drug classes (**G**) and their –log-(p-values) (**H**) as measured with weighted Cox proportional hazards (coxph from the R survival package).

**Table 1.** Hazard ratios and confidence intervals for 6 drug classes across 3 neurodegenerative indications. ALS = Amyotrophic lateral sclerosis, MG = myasthenia gravis, and PD = Parkinson’s disease. CI = 95% confidence interval.

| *Pathway Name* | ALS - HR | ALS - CI low | ALS- CI high | MG - HR | MG - CI low | MG - CI high | PD - HR | PD - CI low | PD - CI high |
| --- | --- | --- | --- | --- | --- | --- | --- | --- | --- |
| CXCR5 | 1·151 | 1·021 | 1·298 | 1·143 | 1·052 | 1·241 |  |  |  |
| CXCR3 | 1·124 | 0·995 | 1·269 | 1·321 | 1·223 | 1·427 |  |  |  |
| Chemokine Pathways | 1·123 | 1·018 | 1·239 |  |  |  |  |  |  |
| Neuropeptide Y | 1·088 | 0·985 | 1·203 |  |  |  |  |  |  |
| CNR2 | 1·0·99 | 1·890 | 1·097 |  |  |  |  |  |  |
| Complement System | 1·031 | 0·937 | 1·134 | 1·098 | 1·018 | 1·183 | 0·992 | 0·965 | 1·020 |

| Pathway Name | ALS - HR | ALS - CI low | ALS - CI high | MG - HR | MG - CI low | MG - CI high | PD - HR | PD - CI low | PD - CI high |
| --- | --- | --- | --- | --- | --- | --- | --- | --- | --- |
| CXCR5 | 1.151 | 1.021 | 1.298 | 1.143 | 1.052 | 1.241 |  |  |  |
| CXCR3 | 1.124 | 0.995 | 1.269 | 1.321 | 1.223 | 1.427 |  |  |  |
| Chemokine Pathways | 1.123 | 1.018 | 1.239 |  |  |  |  |  |  |
| Neuropeptide Y | 1.088 | 0.985 | 1.203 |  |  |  |  |  |  |
| CNR2 | 1.0.99 | 1.890 | 1.097 |  |  |  |  |  |  |
| Complement System | 1.031 | 0.937 | 1.134 | 1.098 | 1.018 | 1.183 | 0.992 | 0.965 | 1.020 |

### Network derived drug classes activate complement system proteins

We next sought additional data sources and experiments to better understand possible drug-induced effects on the complement system. We first sought drug-induced gene expression changes from two public data sources, PharmOmics^21^ and LINCS^22^ (*Supplemental methods*) and saw little or ambiguous effects in these data sources (*supplemental results*). We next hypothesized that protein-level changes may better inform the observed effect on overall survival. We investigated complement levels using ELISA, specifically measuring C3, Complement 4 (C4), and factor B levels released from healthy stem cells following exposure to drugs from our network and non-network classes (**Figure 4**). Healthy stem cells were a sufficient system because we started following patients before their ALS diagnosis and this could indicate whether study drugs induced or suppressed complement system proteins. We prioritized compounds for experimental testing based on those that had the highest number of unique patients and total exposure days per patient (reflecting repeat prescription claims). The CS-class drugs, mirtazapine and methylprednisolone increased activation of both C3 and C4, and the CS-class drug, gabapentin, appeared to have no effect on either C3 and C4. Of the non-CS-class drugs tested, cephalexin, amoxicillin, cefuroxime, nitroglycerin, and solatol had no effect on C3 or C4 levels, however, the non-CS-class drugs, famotidine and erythromycin, increased C3 and C4 levels (**Figure 4**). Drugs from both classes seemed to induce Factor B, but to a lesser effect. Some of these effects are supported by additional literature such as different methylprednisolone regiments in myasthenia gravis patients led to an increase in C3 and C4 levels^23^. This increase in C3, C4 and Factor B for the above compounds indicate that these compounds induce further complement activation once complement activation is initiated, since treatment of human iPSC astrocytes alone w/o the A1 cocktail factors did not elicit any complement response (undetectable levels).

**Figure 4.**
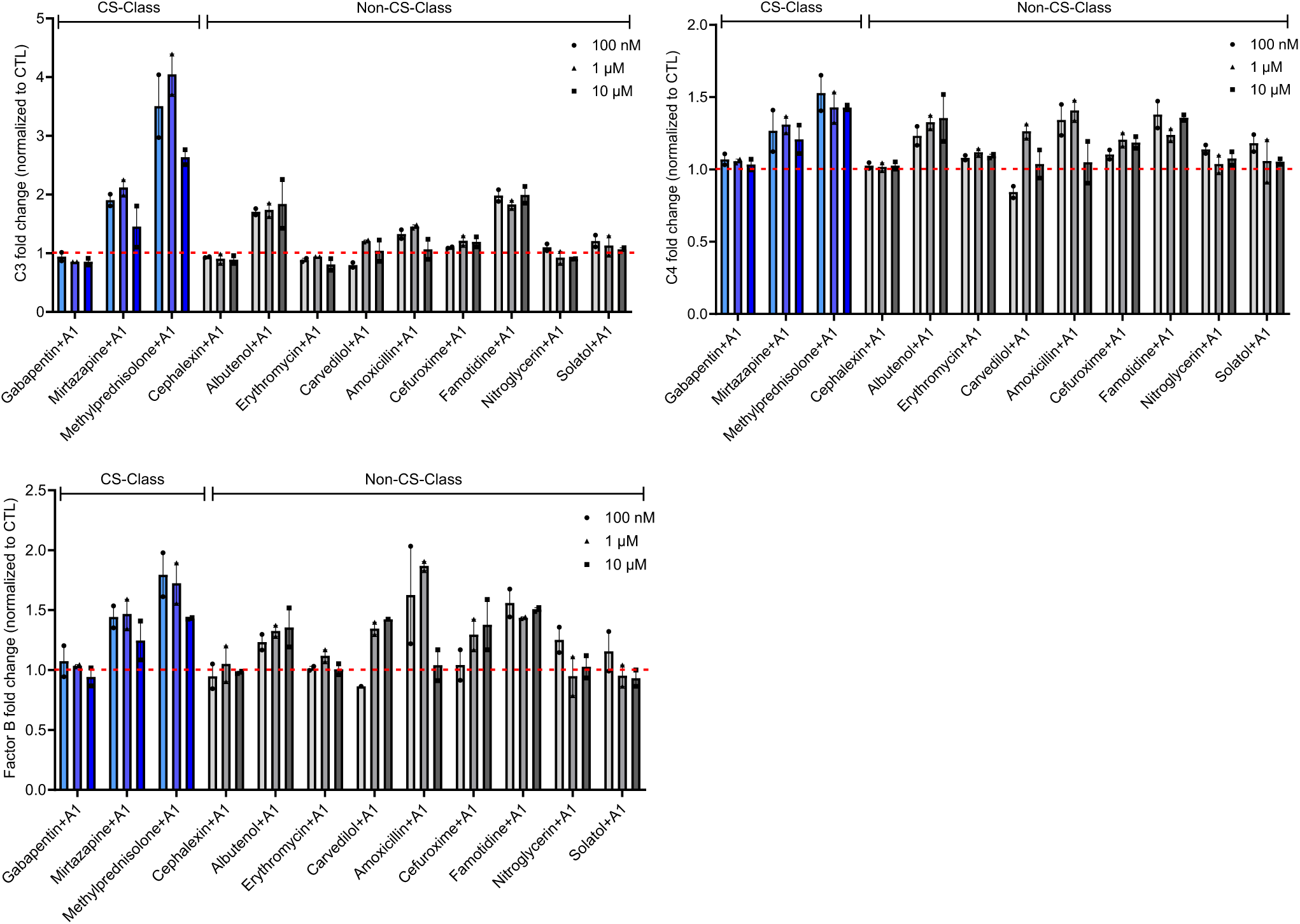
Complement-associated drugs activate multiple components of the complement pathway in ELISA assays. Normalized values (to CTL: A1 cocktail factors) of fold change for the complement factors C3, C4, and Factor B following treatment of human iPSC astrocytes with the A1 cocktail factors and selective classes of compounds (100 nM, 1 µM and 10 µM). Addition of methylprednisolone, mirtazapine and albuterol showed the strongest effect in increasing the complement levels (especially C3).

In our network model, methylprednisolone, gabapentin and mirtazapine have distinct network connections to the complement system proteins, C3, C5, complement C3a receptor (C3AR1), and complement C5a receptor (C5AR1). Specifically, methylprednisolone, gabapentin, and mirtazapine connected through annexin A1 (ANXA1), the adenosine A1 (ADORA1) receptor, and the opioid receptor kappa 1 (OPRK1), respectively (all pathway connections in **Supplemental File 2**). This suggests that alternative pathways could have differential effects on the complement system, but that the observed trends in overall survival might be associated with increased activation of the complement system.

### Several network classes alter overall survival in ALS-patients

We repeated our observational study analysis pipeline for an additional 5 drug classes and ranked their overall effects on survival (**Figure 3B-H**, **Table 1**). We observed a large number of prescription-days per study drug as observed with the complement system study, though, the exact brand names changed per study (**Table S6**, **Figures S9, S11, S13, S15, S17**). Again, we saw low predictive performance of the logistic regression, suggesting good covariate balance (**Table S7**), and similar propensity score plots, suggesting sufficient class balance (**Figures S10, S12, S14, S16, S18**). Across all drug classes, we saw an increased risk for death in the network drug class compared to the non-network classes (**Figure 3G,H**, **Table 1**). The CXCR5 class had the greatest increased risk of death (HR = 1.151, 95% confidence interval (1.021-1.298), p = 0.0214) (**Figure 3B**), and the chemokine class had the next highest risk (HR = 1.123, 95% confidence interval (1.018-1.239), p =0.0202) (**Figure 3C**). The CNR2 class had a marginally decreased, but not significant risk (HR = 0.994, 95% confidence interval (0.890-1.097), p = 0.897), most similar to the value observed for the complement system drug class. All other survival curves provided in **Figures S19-S21**. All of the retained brand names, the number of unique patients with an RX claim, the prescription-days per drug, and patient demographics are included in the extended supplement for each network class comparison: chemokine (**Supplemental File 6**), CXCR3 (**Supplemental File 7**), CXCR5 (**Supplemental File 8**), CNR2 (**Supplemental File 9**), and NPY (**Supplemental File 10**). All hazard ratios also reported in **Supplemental File 16**. We also investigated the extent to which network drug classes shared brand names and observed shared, but distinct brand names in each drug class comparison as measured by Jaccard similarity (**Figure S22C, D,** *Supplemental methods and results*).

### Network-drug classes affect overall survival in multiple neurodegenerative diseases

An intriguing utility for our analysis could be in identification of key nodes of pathophysiology across multiple diseases on the nervous system. We first demonstrated that neurodegenerative diseases shared pathways information using network models (**Figure S23**, *Supplemental methods and results*).

Intriguingly, neurodegenerative diseases share more genes near druggable targets than if comparing all disease genes. We thus repeated measurements for some network classes in multiple disease indications. We specifically considered effects of the complement system in myasthenia gravis and Parkinson’s disease and the CXCR3, and CXCR5 classes in myasthenia gravis. We again assessed prescription days per drug for all studies (**Table S6, Figures S24, S26, S28, S30**), measured low predictive performance of a logistic regression analysis (**Table S7**, and saw similar propensity score plots between target and comparator cohorts (**Figures S25, S27, S29, S31**). Interestingly, we observed a similar trend for myasthenia gravis patients taking the complement system drugs where early survival (before ∼2,000 days) showed separation between the CS-class and non-CS-class drugs, however, the curves crossed-over at later timepoints (∼10,000 days), suggesting a protective effect of complement-associated drugs at early time points. Unlike our analysis in ALS patients, this analysis yielded a slight, but significant increased risk for death (HR 1.098, 95% confidence interval (1.018-1.183), p = 0.015) (**Figure 3D**). This finding combined with our experimental results and the recent approvals of complement inhibitors for myasthenia gravis further suggest that activated complement is detrimental for overall survival in neurodegeneration, but only at early timepoints. The effect of CXCR5 drugs was similar in myasthenia gravis patients relative to ALS patients (HR 1.143, 95% confidence interval (1.052-1.241), p = 0.00155) (**Figure 3E**) and the effect of CXCR3 drugs was greater than in ALS patients (HR 1.321, 95% confidence interval (1.223-1.427), p = 1.47 x 10^-12^) (**Figure 3F**). The effect of complement-system drugs in Parkinson’s patients showed, similar, but starker cross-over effects like we observed in ALS and myasthenia gravis patients. We observed no overall survival benefit or risk (HR 0.992, 95% confidence interval (0.965-1.020), p = 0.579) (**Figure S32**). Like before, all of the retained brand names, the number of unique patients with an RX claim, the prescription-days per drug, and patient demographics are included in the extended supplement for each network class comparison: CXCR5 in myasthenia gravis (**Supplemental File 12**), CXCR3 in myasthenia gravis (**Supplemental File 13**), complement system in myasthenia gravis (**Supplemental File 14**), and complement system in Parkinson’s (**Supplemental File 15**).

## Discussion

We presented an approach for using proteins downstream of druggable targets to define new drug classes and tested the effects of these drug classes on overall survival in ALS and two additional neurodegenerative diseases. We used observational studies as exploratory analyses, and while they would require further, dedicated clinical investigations to confirm their effects, they are compelling for their ability to recover known effects – specifically the effect of complement activation as detrimental to survival in ALS, myasthenia gravis, and Parkinson’s disease. Our preliminary experimental data combined with recent approvals of complement inhibitors for myasthenia gravis patients (**Table S4**) further supports the potential of complement inhibitors in ALS. Overall, this suggests a novel and impactful approach for identifying disease-drivers in rare and difficult-to-treat diseases and may provide valuable insights to therapeutic target selection.

Our results are further compelling for their utility in difficult-to-treat populations. ALS is insufficiently explained by genetics, with less than 10% of familial and sporadic ALS cases explained by genetic drivers^9,10^. Without sufficient molecular targets, novel approaches are desperately needed to identify treatment strategies for the disease. Further, the ALS population is relatively small and is typically insufficient for observational studies. Indeed, through a related project, the Veterans Affairs dataset, which contained nearly 20,000 ALS patients, is the largest repository of historical ALS patient data and is nearly double the size of the cohort in this study^24^. Yet, we still resolved statistically-significant clinical effects by emphasizing proteins downstream of druggable targets, which allowed the inclusion of more patients.

While we believe our paradigm can greatly enhance model-informed drug development efforts, further work could overcome limitations in our current study. For instance, we had insufficient network connections to all possible druggable targets and were limited to deriving network classes for proteins that were “close” to approved targets. Our network drug classes also shared many approved drugs indicating that our current interaction network was only sufficiently connected to some pathways. However, the differences in network phenotypes effectively separated drugs to discern relatively stronger effects, such as the increased risks associated with CXCR5 networks. This emphasizes the importance of deriving high-quality, disease pathway phenotypes. We also tested these network classes in a single patient population and further testing using data from additional health systems could elevate the impact of these findings. Indeed, previous meta-analyses across healthcare systems have successfully discovered drug-class effects on cardiovascular disease outcomes^25^. Further, an intriguing opportunity, or possible challenge, of historical data analysis is how to interpret the outcomes in the context of EHR timescales and understand their relevance to dedicated clinical investigations. We followed patients for nearly two decades, when most clinical trials are months to years in duration. Of the four FDA-approved therapies for ALS, the riluzole trials measured outcomes at 12 months^26^ or 18 months follow-up^27^ and the trials for edavarone, phenylbutyrate-taurursodiol (PB-TUDCA), and torfersen measured changes in the revised ALS functional rating scale^28^ at shorter durations – ranging from 24-28 weeks^9,29–32^. Our measured differences in proportions of surviving patients at short durations (∼2 years) are similar to effect sizes in the riluzole trials, suggesting that future observational studies, which leverage drug network classes, could shorten observation times to better anticipate effect sizes measurable in the duration of a clinical study.

Our discovery of disease drivers complements an evolving landscape of shared molecular underpinnings of clinically-distinct diseases. Specifically, we predicted that activated complement would increase risk for death across multiple disease indications. Our pathways analysis of shared dysregulation is supported by other findings, specifically, Arneson et al^33^ noted that when disease genes and proteins weren’t shared across neurodegenerative diseases, gene and protein functional information was shared, further emphasizing shared dysregulation across neurodegenerative diseases. Separate investigations of post-mortem brain samples further corroborated shared gene-level changes across patients with four neurodegenerative diseases (Alzheimer’s Disease, Parkinson’s disease, Huntington’s disease, and ALS) and discovered that shared genes were involved in functional processes such as inflammation, mitochondrial dysfunction, and oxidative stress^33,34^. Additionally, multiple canonical inflammatory reactivity inducers, including complement subcomponent C1q, interleukin 1 alpha (IL-1⍺), and tumor necrosis factor (TNF), convert astrocytes into A1 reactive astrocytes^35^, and these proinflammatory, reactive astrocytes secrete multiple complement factors, reduce synaptic and neuronal connectivity, and are linked extensively to neurodegenerative disease^36^. Taken together, this further supports the possibility of shared molecular changes in these diseases, though, further experimental validation would be required to confirm this general finding.

Our approach motivates extending the integration of clinical health data into Model Informed Drug Development (MIDD). Already, the FDA has provided guidance about the use of real-world data (RWD) for generating real world evidence (RWE) from sources other than randomization, controlled trials (RCTs)(**Table S9**). Indeed, many have generated RWE to support regulatory decision making including accelerated approvals, learning intrinsic factors, and optimizing dosing (**Table S9**). Yet these studies did not leverage RWD for novel target support.

Other sources have identified novel targets from RWD, although, they have focused directly on target-driven effects instead of downstream effects. Genetic data repositories have helped identify lead candidates from patient data, such as in the case of Proprotein convertase subtilisin/kexin type 9 (PCSK9) inhibitors where inactivating and gain-of-function mutants were associated with elevated or reduced cholesterol, respectively^37,38^. Mendelian randomization is another method of inferring drug-target effects in new populations^39^, but not every suitable target has sufficient genetic connections to support later therapeutic development. Additionally, analysis of PheWAS and expression quantitative trait loci (eQTLs) have also coupled gene variation with observable clinical effects, especially side-effects^40,41^. In contrast to our approach, these approaches overlook downstream effects and require sufficient genetic variation in patient populations to advance therapeutic discovery.

For the most part, prior work combining molecular models of drug effects with patient data have been limited to drug repurposing studies, and we believe our approach greatly expands the possibilities for learning promising therapeutic targets from real-world patient data. Knowing that electronic health records will be increasingly useful to support new target identification, we anticipate that network medicine will extend capabilities beyond target-centric approaches.

## Study Highlights

Currently, the impact of real-world data (RWD) on druggable targets with translational impact is limited to drug-repurposing. However, we hypothesized that network models of drug-induced signaling could elevate the impact of RWD for identifying novel drug targets. This study addressed the feasibility of that approach by simulating the effects of novel targets and testing these effects in neuroinflammatory conditions. We demonstrated that drugs that alter complement activation have deleterious effects on overall survival and these results are consistent with effects of recently FDA-approved drugs, although none of our patients were taking them. We believe this paradigm will change translational science in how RWD may be used to identify novel, druggable targets.

## Supporting information

Supplemental methods, results, tables, figures

## Acknowledgments

The authors would like to acknowledge the Sanofi real world data team, and especially Thomas Verrier, the RWD-Platform Support team for their invaluable support navigating data access, without which, the study could not be completed.

## Contribution statement

J.L.W., M.A., J.Y.H., G.D., A.S.B., F.F., D.O. wrote the manuscript. J.L.W., J.Y.H, G.D., A.S.B., D.O. designed the research. J.L.W., J.Y.H., M.A., G.D. performed the research. J.L.W., M.A., G.D. analyzed the data. A.S., G.G., T.H., F.F., MC. M. contributed new reagents/analytical tools.

## Supplementary Material

Supplemental File 1 – extended methods, results, Tables S1-8, Figures S1-32.

Supplemental File 2 – ALS-PathFX pathways, all modeling results.

Supplemental File 3 – All network-protein derived drug classes.

Supplemental File 4 – Drug brand names, prescription days, patient demographic data, and covariates for the study of ALS patients and the CS-class.

Supplemental File 5 – All diagnosis codes used to select patients.

Supplemental File 6 – Drug brand names, prescription days, patient demographic data, and covariates for the study of ALS patients and the CXC-class.

Supplemental File 7 – Drug brand names, prescription days, patient demographic data, and covariates for the study of ALS patients and the CXCR3-class.

Supplemental File 8 – Drug brand names, prescription days, patient demographic data, and covariates for the study of ALS patients and the CXCR5-class.

Supplemental File 9 – Drug brand names, prescription days, patient demographic data, and covariates for the study of ALS patients and the CNR2-class.

Supplemental File 10 – Drug brand names, prescription days, patient demographic data, and covariates for the study of ALS patients and the NPY-class.

Supplemental File 11 – PathFX predictions for multiple neurodegenerative diseases.

Supplemental File 12 – Drug brand names, prescription days, patient demographic data, and covariates for the study of MG patients and the CXCR5-class.

Supplemental File 13 – Drug brand names, prescription days, patient demographic data, and covariates for the study of MG patients and the CXCR3-class.

Supplemental File 14 – Drug brand names, prescription days, patient demographic data, and covariates for the study of MG patients and the CS-class.

Supplemental File 15 – Drug brand names, prescription days, patient demographic data, and covariates for the study of PD patients and the CS-class.

Supplemental File 16 – Table of hazard ratios and confidence intervals for all studies.

