## Supplemental methods, results, tables, figures for "Learning good therapeutic targets in ALS, neurodegeneration, using observational studies"

*Curating ALS-associated neurogenerative disease pathways*

We curated multiple datasets from patient data, public databases, and consortia to create our custom platform. Here, we describe data processing for each source.

a. TargetALS – We obtained raw fastq RNA-Seq data files (1,440 total) from the NYGC ALS Consortium (Target ALS Release July 2022) (https://www.targetals.org/research/resources-for-scientists/resource-genomic-data-sets/). These fastq files were generated using human postmortem tissue samples from the Target ALS postmortem tissue core and processed for RNA-Seq as previously described1. Qiagen processed the raw fastq files in OmicSoft ArrayStudio RNA-Seq analysis pipeline (version 11.7.3.3). In brief, using the fastq files, the pipeline performed quality control and alignment to the Genome Reference Consortium Human Build 38 (GRCh38, GPL16791) using the proprietary OmicSoft Aligner2. After alignment, we determined the gene level RPKM/FPKM/counts using the EM algorithm in ArrayStudio as described previously3. Finally, we calculated pairwise differentially expressed genes using the DESeq2 v1.10.14 in ArrayStudio comparing tissue-specific ALS patients vs. non-neurological controls. We excluded ALS patients with multiple neurological conditions from pairwise analysis and we considered differentially expressed genes (DEGs) as significant with a padj $< 0.05$.
b. We included 13 pathways derived from differentially-expressed genes from RNA-seq in multiple tissues from the TargetALS consortium. Specifically, we assessed genes with log(fold-changes) > 1.0, or log(fold-changes) > 1.5 in several tissues. Because PathFX required 25 genes to define a pathway phenotype, we only retained pathways for the 13 samples listed in **Table S1**. In total, this yielded 16,570 gene-phenotype relationships from 13 disease pathways and 8,081 unique genes. While all pathways and their sizes (i.e., gene list lengths) are included in **Supplemental File 2**, we've highlighted pathway names extracted from Target ALS data to demonstrate the tissues used (**Table S1**).

| Tissue | Gene List derived |
|---|---|
| Cerebellum | Target ALS log2fc>1 Cerebellum |
| Spinal Cord | Target ALS log2fc>1 SpinalCord |
| Spinal Cord | Target ALS log2fc>1 SpinalCordCervical |
| Spinal Cord | Target ALS log2fc>1 SpinalCordLumbar |
| Cerebellum | Target ALS log2fc>log1.5 Cerebellum |
| Cortex | Target ALS log2fc>log1.5 FrontalCortex |
| Cortex | Target ALS log2fc>log1.5 PrimaryMotorCortex |
| Cortex | Target ALS log2fc>log1.5 PrimaryMotorCortexLateral |
| Cortex | Target ALS log2fc>log1.5 PrimaryMotorCortexMedial |
| Spinal Cord | Target ALS log2fc>log1.5 SpinalCord |
| Spinal Cord | Target ALS log2fc>log1.5 SpinalCordCervical |
| Spinal Cord | Target ALS log2fc>log1.5 SpinalCordLumbar |
| Spinal Cord | Target ALS log2fc>log1.5 SpinalCordThoracic |

**Table S1.** Target ALS pathways

c. PathFX v2 – We searched the PathFX v2 database (released with [1]) for ALS and neurodegenerative related phenotypes. Specifically, we searched for phenotypes with 'Lou Gehrig', 'amyotrophic', or 'sclerosis' in their names, manually removed any phenotypes that were erroneously captured (e.g., 'Arteriosclerosis'), and then further removed phenotypes with fewer than 25 genes because we considered these as insufficiently connected to do pathways analysis based on prior findings[2]. This yielded 60 phenotypes. We additionally added 1,000 randomly-sampled phenotypes from the PathFXv2 database because we previously discovered that the algorithm needed a sufficient number of total pathways to correctly calibrate the expected association of drug to a pathway phenotype (*unpublished*). In total, this yielded 170,155 gene-phenotype relationships across 1,060 disease pathways and 19,406 unique genes.

d. We incorporated novel sequencing datasets collected from multiple cell and tissue types relevant to ALS [3]. They included 11 snRNAseq pseudobulk data sets from multiple cell lines: OPC, astrocytes, endothelial cells, ependymal cells, fibroblasts, lymphocytes, microglia, neuron, oligodendrocytes, and pericytes. We also included a disease pathway assessing snRNAseq changes across all cell lines, a bulk, all tissue RNAseq in ALS compared to control patients, and a proteomics dataset comparing abundance in “high” vs “low” ALS patients. In total, this yielded 5,947 gene-phenotype relationships across 14 disease pathways and 3,399 unique genes.

e. Reactive astrocytes – we incorporated differential gene expression from reactive astrocytes. This yielded 1,132 gene-phenotype relationships to one disease pathway.

f. Cerebral spinal fluid mass spec – we also used protein abundance data captured from mass spectrometry of patient cerebral spinal fluid. This yielded 50 gene-phenotype relationships associated with one disease pathway.

g. Microglia – We also included gene expression changes captured in microglia using single nucleus sequencing data. From this data, we derived eight pathways using multiple thresholds for the log(fold-change): 1.5, 1.7, 1.9, 2.1, 2.3, 2.5, 2.7, and 2.9. This yielded 1,398 gene-phenotype relationships across 8 disease pathways and 360 unique genes.

*Generating PathFX-ALS*

We constructed a custom version of PathFX, (ALS-PathFX), to discover drug networks with associations to omics data relevant to amyotrophic lateral sclerosis (ALS) model systems and patient samples. PathFX[4] is a network-based platform designed to predict drug effects by analyzing the interactions between drugs and their target proteins within biological pathways. It utilizes protein-protein interaction networks and can integrate various omics data to identify potential therapeutic targets and predict the downstream effects of drug actions. As published in[4], PathFX incorporates lists of genes and/or proteins when assessing phenotypes associated with drug networks. Generating a new PathFX version required repeating permutation tests that mitigated biases associated with network analysis methods. In the PathFX-v1[4] and PathFX-v2 (released with [1]), algorithm generation, we used two bias reduction techniques: "interaction specificity" analysis which reduced the likelihood of high-degree network nodes from being over-represented in drug networks and a second process that minimized annotation bias, or the tendency for pathways with larger gene lists to be included in more drug networks. For this version, we used the same interaction network as released with PathFX-v2 and did not repeat interaction specificity analysis. However, because we updated the list of disease pathways and their associated genes and proteins, we measured pathway associations to 100 random networks to estimate the expected chance of association. ALS-PathFX used expected associations to remove phenotypes from a real drug's network that were less significantly associated than the median of the 100 random networks. This threshold was set per phenotype, because each phenotype had a unique set and number of associated genes/proteins.

*Selecting patient cohorts for observational studies*

We first identified patients diagnosed with ALS, or later with additional neurodegenerative diseases, using several diagnosis codes (**Supplemental File 5**). We further divided these patients into target or comparator cohorts based on whether they had a prescription for one of the active ingredients in the network or non-network drug classes (described above). We removed any patients in both cohorts. For all patients, cohort entry was defined by the start of the active ingredient (measured by first prescription fill-date associated with an RX claim) and cohort exit was considered as the first day of the month of death (the data table only recorded month and year of death).

*Propensity score analysis using logistic regression of patient features*

Data-driven propensity score analysis is a statistical technique used to reduce bias in observational studies by creating comparable groups based on their likelihood of receiving the treatment. This method has demonstrated power in reducing confounding in observational studies[5–7]. It further reduces the need to specify known confounders and allows investigation of any patient features that could explain assignment to the target or comparator groups.

In this study, we utilized logistic regression to estimate the probability (propensity score) that a patient receives the treatment (i.e., one of the network class drugs) based on multiple, possible covariates.

For all study patients, we specifically assessed all non-ALS diagnosis codes (or non-myasthenia gravis or non-Parkinson's codes in later studies), and all non-study prescription drugs as sufficient to describe the patient's underlying biology. We counted all non-ALS diagnosis codes that were within 365 days prior to the start of a study drug and retained those shared by at least 500 patients, and counted all non-study drugs that were prescribed within 365 days prior to the start of a study drug and kept those associated with at least 100 patients (this thresholding changed for non-ALS disease groups because the patient cohorts were larger and low-frequency codes were less likely to predict cohort assignment). We also added year of birth, sex, race, and ethnicity when available. We created a pivot table for the top diagnosis and prescription features, and coded sex as 1(0) for male(female), race/ethnicity as 'Black':1, 'Asian':2, 'White':3, and 'Hispanic':4.

We conducted logistic regression analysis using the sklearn module in python to predict assignment to the target group using the above prioritized patient features. When logistic regression was poorly predictive of treatment assignment, we considered that the cohorts were well balanced for further observational study. We also further inspected the top/bottom regression coefficients to assess if any patient features were associated with either drug class (**Supplemental File 4**).

We repeated the same confounding procedure for the other five drug classes and additional two neurodegenerative diseases and discovered good covariate balance across cohorts. We further reported the top/bottom regression coefficients for all studies (**Supplemental Files 6-10, 12-15**).

*Effect estimation*

We used the survival package in R to perform a weighted effects estimate using predicted probabilities generated from the above logistic regression, and specifically used the 'coxph' function in the 'survival' package and provided weight values to the method using the inverse of the propensity score. All values reported in **Table 1** are also recorded in **Supplemental File 16**.

*Assessing top/bottom gene expression in PharmOmics*

We assessed complement proteins in gene signature data from the PharmOmics dataset[8]. PharmOmics is a large meta-database that merged several gene-expression datasets selected from multiple platforms after treatment of drugs in various tissues and in multiple species (mouse, rat, and human). From their publication, we quote their description: "A total of 941 drugs, including 766 FDA approved drugs from KEGG, FDA, European Medical Agency, and Japanese Pharmaceuticals and Medical Devices Agency, and 175 chemicals from TG-GATEs and DrugMatrix were queried against GEO, ArrayExpress, TG-GATEs, and DrugMatrix to identify datasets"[8] The data is provided as a large datamatrix in which drugs are linked to genes that are upregulated ("top") or downregulated ("bottom") in any GEO dataset with that drug treatment. Using only the human data, we counted if complement proteins were in the top/bottom list for the CS-class and non-CS-class drugs using the pandas module in Python.

*Gene expression from the LINCS dataset*

We accessed the level five z-score normalized gene expression data from LINCS[9]. We specifically searched for drug names that matched branded names in our analysis. LINCS contains several cell lines, however, we restricted our search to cell lines that could be relevant for ALS. After filtering for the intersection of our drug names, we retained 456 drug-cell line gene expression profiles, associated with 354 unique drugs and 2 unique cell lines ('NPC' and 'NEU' cell lines which are both described as 'normal stem cell sample' from the 'central_nervous_system', however NPC is aliased as 'NEURAL PROGENITORS' and 'NEU' is aliased as 'NEURONS' in the LINCS documentation). We then used matplotlib in python to plot the distribution of gene expression scores for eight of the nine complement system proteins (expression data was not available C1QC).

*Stimulation and drug treatment, compounds used*

| Network Class | Drugs | Company | Part Number | MW | Solubility |
|---|---|---|---|---|---|
| CS-class (predicted to affect complement system) | Gabepentin | Fisher Scientific | 5085060001 (10 mg) | 171.24 | Water |
| | Methylprednisolone | Fisher Scientific | 481950 (50 mg) | 374.47 | Water (120 mg/L); DMSO:PBS PH:7.2 (1:1) |
| | Mirtazapine | Fisher Scientific | 201850 (50 mg) | 265.36 | DMSO |
| Non-CS-class (not predicted to affect complement system) | Albuterol or Salbutamol hemisufate | Fisher Scientific | 0634250 (250 mg) | 337.4 | 1M NaOH (50 mg/mL); water up to 100mM |
| | Amoxicillin | Fisher Scientific | AC455140050 (5 g) | 365.404 | Water (1 mg/Ml) |
| | Carvedilol | Fisher Scientific | 268550 (50 mg) | 406.5 | DMSO |
| | Cefuroxime (sodium salt) | Fisher Scientific | 111014629 | 446.37 | Water (50 mg/mL) |
| | Cephalexin | Fisher Scientific | ICN15058525 (25 g) | 365.4 | Water (1-2 mg/mL) |
| | Erythromycin | Fisher Scientific | ICN19019701 (1 g) | 733.94 | Water (2 mg/mL); DMSO |
| | Famotidine | Sigma Aldrich | 1269200 (125 mg) | 337.45 | DMF (Dimethylformamide): freely soluble) |
| | Nitroglycerin (in propylene glycol) | Sigma Aldrich | 1466506-5 AMP | 227.09 | In solution |
| | Solatol (hydrochloride) | Fisher Scientific | 95250 | 308.82 | Water (50 mM) |

**Table S2.** Drugs tested in ELISA by predicted network class

*Analysis of co-occurring diseases in drug networks*

We ran PathFXv2 for all approved drugs in DrugBank and assessed drug associations to several neurodegenerative phenotypes (**Table S3**). We assessed PathFX phenotypes associated to ALS, multiple sclerosis, Alzheimer's disease, stroke, myasthenia gravis, and Parkinson's Disease. For each disease, we found multiple relevant PathFX phenotypes (**Table S3**). We counted how often one of the disease-area PathFX phenotypes co-occurred in drug networks and used Jaccard similarity to compare all six diseases. We plotted a co-occurrence heatmap using seaborn in python. Code for this analysis is provided in our GitHub.

| Disease Group | Phenotype | Number of drugs |
|---|---|---|
| AZ | Alzheimer disease | 1012 |
| AZ | Familial Alzheimer Disease (FAD) | 442 |
| AZ | Alzheimer Disease, Late Onset | 336 |
| AZ | Alzheimer's Disease, Focal Onset | 266 |
| AZ | Alzheimer Disease, Early Onset | 170 |
| AZ | Alzheimer Disease | 99 |
| AZ | ALZHEIMER DISEASE 5 | 13 |
| ALS | Amyotrophic lateral sclerosis | 58 |
| ALS | Amyotrophic Lateral Sclerosis, Sporadic | 6 |
| ALS | Amyotrophic lateral sclerosis type 1 | 4 |
| STK | Cerebral Infarction | 549 |
| STK | Ischemic stroke | 329 |
| MS | Multiple Sclerosis | 534 |
| MS | Multiple Sclerosis, Acute Fulminating | 7 |
| MS | Multiple Sclerosis, Relapsing-Remitting | 1 |
| MYGR | Myasthenia Gravis | 376 |
| PD | Parkinson disease | 421 |
| PD | Parkinson disease 2 | 39 |
| PD | Parkinsonian Disorders | 25 |
| PD | Parkinson disease, late-onset | 12 |

**Table S3**. Neurodegenerative diseases are associated with multiple drug networks

### Supplemental Results

*ALS-PathFX predicted drug network associations to ALS through distinct proteins*

Like acetophenazine, we considered all ALS-PathFX predictions to all neurodegenerative phenotypes and across multiple network protein drug classes. We additionally plotted all network proteins for all approved drugs and highlighted protein classes considered for later discovery (**Figure S1,** all ALS-PathFX predictions in **Supplemental File 2**). Notably, when modeling drugs based on network proteins, drugs clustered distinctly from anatomical therapeutic chemical (ATC) codes, a commonly-used drug classification system based on intended therapeutic use (**Figure S1A**), suggesting that network analysis can uncover similarities across drugs with different therapeutic use cases. Further, looking at level 1 ATC codes, which represent a drug's primary intend-to-treat anatomical system, our analysis predicted that drugs from a broad range of therapeutic uses would have network connections to ALS and neurodegenerative phenotypes (**Figure S1B**).

We were first drawn to the complement system proteins because extensive literature supported dysregulation of these system proteins in neurodegenerative disease and complement inhibitors have been successful in other disease applications (**Table S4**). Additionally, in the ALS-PathFX model, a network protein is predicted to be perturbed by the drug. However, the prediction doesn't describe whether the drug activates or inhibits the protein. In this scenario, we didn't yet know if the complement system is activated or not by the associated drugs. As such, we hypothesized that complement-associated drugs altered the complement system and that these drugs would have distinct clinical effects from drugs without complement system proteins in their networks.

The complement system proteins (we considered nine proteins in total, **Table S5, Figure S1C**) were discovered in 1,945 drug-phenotype relationships across 17 unique disease pathways in the networks for 312 approved active ingredients. Within these drugs, their networks contained distinct associations to each of the complement system proteins (**Figure S1D**). Additionally, 551 active ingredients had networks associated with at least one of the 17 neurodegenerative disease pathways, but did not contain any complement system proteins in their networks (**Figure S1C, 3D**). We considered this our first network-drug class, where the complement-system drugs are represented in light blue (conceptual in **Figure 1**, drug network data in **Figures S1C,D**) and the non-complement associated drugs were the “non-CS” drug class and are represented in light grey (conceptual in **Figure 1**, drug network data in **Figures S1C,D**).

To identify network drug classes generally, we next counted how often a network protein (or group of proteins) connected a drug to an ALS or neurodegenerative disease pathway. In addition to the complement system components, we discovered chemokine receptors, and neuropeptide proteins among the most associated (**Table S5,** all other classes in **Supplemental File 3**) and we reclassified drugs if their networks included the network protein or not. In the case of the complement system, chemokine systems, and neuropeptide Y systems (NPY) we used a list of proteins to define the new classes (**Table S5**. Compared to the complement system, the NPY class was discovered in 2,242 drug-phenotype relationships, across 13 unique phenotypes, and was associated with 305 unique approved drugs (**Figure S1F**). We also plotted all network proteins for the chemokine (**Figure S2**), Cannabinoid Receptor 2 (CNR2) (**Figure S3**), C-X-C Motif Chemokine Receptor 3 (CXCR3) (**Figure S4**), and C-X-C Motif Chemokine Receptor 5 (CXCR5) (**Figure S5**) classes. For many network-associated proteins, they occurred infrequently in drug networks and generated relatively small drug classes (**Supplemental File 3**).

Because we wanted to conduct target-comparator analyses in the electronic health record, we generated a list of comparator drugs based on the exclusion of the network protein, as described in the conceptual motivation. The previously-described 274 active ingredients without complement system proteins in their networks became the comparator drug class for the complement-class drugs. As one other example, we repeated this process for the NPY drug class, and discovered 587 unique approved drugs associated with networks connected to the same disease pathways, but did not have NPY, or its receptors, in their drug network (**Table S5**). Taken together, ALS-PathFX identified thousands of predicted effects for marketed drugs on ALS and neurodegenerative disease pathways and these predicted effects were associated with distinct non-drug-target, network proteins. We observed that many network classes yielded imbalanced or empty comparator cohorts (**Supplemental File 3**), making them impractical to study in EHR data; having a reasonably similar number of approved active ingredients in target-comparator drug classes would be required for later analysis in electronic health record data, indicating that not all potential targets could be tested in our pipeline.

*A paradigm for testing network drug class effects in retrospective observational studies*

We first filtered all patient entries for those with a diagnosis code of “Amyotrophic Lateral Sclerosis” (all diagnosis codes for the study included in **Supplemental File 5**) and retained patients with an exposure to one of the complement class drugs (**Table S6**, “Number of Approved Drugs”, “CS Class”) or one of the drugs with similar network phenotypes but no complement system proteins in their networks (**Table S5**, “Number of approved drugs w/o the protein or group”, “Non-CS Class”). In our study, cohort entry was defined as the time of the first prescribed CS/non-CS class drug; this included some patients ahead of their ALS diagnosis. We removed patients who were in both the CS and Non-CS Class. We next assessed the total number of exposure days per prescription per patient (**Figure S6A**, number of patients per class drug included in **Supplemental File 4**). Patient prescriptions are an imperfect means for estimating drug exposures, but they are a useful proxy for exploratory analysis. Generally, we observed relatively high total exposure days (summed total of all prescription days per patient), suggesting that patients were taking the drug and refilling prescriptions. In the end, the target group contained 40 prescribed brand names (not all modeled active ingredients were prescribed to ALS patients) prescribed to 1,312 unique patients, and the comparator group contained 38 unique brand names prescribed to 2,243 unique patients (**Table S6**, and **Supplemental File 4**).

*Propensity-score analysis confirms sufficient covariate balance*

We next conducted large-scale propensity scoring to control for possible confounding factors. Large-scale propensity analysis uses additional patient information and logistic regression to predict the likelihood of being prescribed the “treatment”; these models have led to unbiased assessment of treatment effects in the electronic health record[17–20]. For our model, we considered all non-ALS diagnosis codes, prescription claims not assigned to either the CS/non-CS drug classes, and the patient demographics, date of birth, race, and sex (all covariates listed in **Supplemental File 4**). We used logistic regression modeling to predict treatment assignment and found poor predictability (mean-squared error 0.32, coefficient of determination=-0.37, **Table S7**). Because patient features were not predictive of treatment assignment, this suggested low confounding between patients in the target or comparator groups. Essentially, patients with similar features were represented in the treatment and comparator groups (**Figure S6B**) (regression coefficients in **Supplemental File 4**). Additionally, we explored time-to-death in both cohorts and observed no enrichment for “fast” or “slow” responders in either cohort (**Figure S7**). We explored the demographics of this patient group and discovered that the population was primarily white (**Figure S6C**), with relatively similar numbers of males and females (**Figure S6D**), and the patients’ year-of-birth (YOB) was similarly distributed between both classes (**Figure S7**) (non-CS-class YOB median=1945, min=1912, max=2011; CS-class YOB median=1948, min=1918, max=1991) (**Supplemental File 4**).

*Complement-associated drugs show no changes in gene expression*

To estimate this effect, we first used the PharmOmics dataset[8], which measured gene expression changes after drug exposure in multiple animal models and tissues, as gathered through a meta-analysis of several gene expression datasets. Although the dataset doesn’t emphasize samples relevant to ALS, it is a large repository that was likely to contain data for many of the drugs modeled in ALS-PathFX. The dataset included gene names if they were in the top or bottom differentially-expressed genes per drug and so we counted how often a complement system protein occurred in the top or bottom expressed genes for network and non-network class drugs. We discovered that 65 and 72 of the CS-class and non-CS-class drugs were in PharmOmics, respectively, and that complement proteins were in the “top” and “bottom” for several of the drugs. However, we found no substantial patterns in expression (**Table S8**). The CS-class drugs had slightly more instances of C5 in the “top” genes (five CS-class drugs compared to one non-CS-class drug) where the non-CS-class had slightly more instances of C5 in the “bottom” genes list (eight non-CS-class drugs compared to four CS-class drugs). However, this was a limited dataset, with relatively few of our study drugs, and our observations are without statistical testing. We additionally turned to the LINCS dataset[9] which contained continuous differential expression data for several drug perturbations across multiple cell lines. We plotted the gene expression for eight of the nine complement system proteins (C1QC was not in the dataset) using two neuronal cell lines (these cell lines contained data for 354 of the CS and non-CS class drugs). Again, we observed no striking differences in the expression of any complement system protein between the network drug classes (**Figure S8**).

*Phenotypic differences drive differences in network-drug classes*

Indeed, the target drug classes at the network level shared many approved drugs, with the chemokine, CNR2, CXCR3, and CXCR5 target classes all sharing the same drugs (**Figure S22A**). However, the comparator drug classes were relatively unique because these were derived from similar pathway phenotypes (**Figure S22B**) and no two comparator classes were the same. Looking at the disease pathways associated with each class, we observed differences in the disease phenotypes (**Figure S22E, Supplemental File 11**). For instance, when comparing the complement system and chemokine drug classes**,** the former is uniquely associated with “pericytes snRNAseq”, “Target ALS RNAseq from the cervical spinal cord,” and “oligodendrocyte snRNAseq" and that latter is uniquely associated with eight disease pathways including “Amyotrophic Lateral Sclerosis, Sporadic”, “cerebral spinal proteomics”, and “Target ALS RNAseq from the entire spinal cord” (**Supplemental File 11**). While outside the scope of this work, this suggests potential tissue- and cell- specific mechanisms for differences across the network protein classes.

*Neurodegenerative diseases shared molecular pathways*

Other studies have shown a clear link between the molecular and genetic features of several neurodegenerative diseases [10,11], with some studies emphasizing shared gene and protein functional information in cells and post-mortem samples[10,12]. Additional studies classified ALS on a frontotemporal dementia -motor neuron disease (MND) continuum[13,14], giving the disease characteristics of other neurodegenerative disease. This suggested that our findings in ALS may be generalized to other neurodegenerative indications, especially if drug networks contained associations to additional neurodegenerative disease pathways. Additionally, other neurodegenerative patient populations were larger and could further test our earlier results.

We first assessed the PathFXv2 database (because this database was larger than our ALS-PathFX database) for phenotypes related to Alzheimer’s disease, Parkinson’s disease, multiple sclerosis, myasthenia gravis, and stroke. Using a simple string-matching approach, we discovered 45 phenotypes related to these indications, with sufficient connections to the interactome for PathFX modeling (175 other phenotypes were discarded because they were insufficiently connected to the interactome). We again assessed PathFX network associations to the 45 phenotypes and analyzed their shared pathway information. This analysis discovered 4,699 drug-disease associations between 1,257 drugs and 20 disease pathways (**Supplemental File 11**). The phenotype, "Alzheimer disease" was associated to the most drug networks (1,012 unique drugs), and the phenotype "Multiple sclerosis, relapsing-remitting" was associated to the fewest drug networks (one drug) (all phenotypes and number of associated drugs listed in **Table S3**).

For instance, the mirtazapine drug network was associated with "Alzheimer disease", "Parkinson disease", and "Multiple Sclerosis" (**Figure S23A**). These associations were also driven by the six network protein classes. As another example, "Alzheimer disease" and "Multiple Sclerosis" also co-occurred in the network for tasonermin (**Figure S23B**), but this network was not connected through any of the network class proteins and would have only been considered in the comparator drug class. These networks also demonstrate that although "multiple sclerosis" and "Alzheimer's disease" co-occurred in two drug networks, their network genes were distinct (i.e., tasonermin's network proteins – TRADD, TRAF2, BIRC2, BIRC3, and RIPK1 – were not used to predict mirtazapine's associations to the same phenotypes).

We next grouped all phenotypes based on their related diseases (e.g., "cerebral infarction" and "ischemic stroke" were considered related to "stroke") and assessed drugs shared between disease groups using Jaccard similarity (**Figure S23C**). Myasthenia gravis and multiple sclerosis phenotypes co-occurred most often, with stroke and Alzheimer's disease co-occurring the second most often. Parkinson's disease and ALS were the least frequently co-occurring with the other neurodegenerative disease indications. Recently, we discovered that drug networks with similar predicted phenotypes shared a greater proportion of disease proteins than the diseases individually (unpublished), suggesting that drug networks converge on shared subsets of disease pathways. We last quantified the drug-network-associated genes for co-occurring neurodegenerative diseases (**Figure S23D**) and genes shared among the disease pathways (**Figure S23E**). Interestingly, like our unpublished finding, the network-associated disease genes were more similar than all pathway genes. For instance, in **Figure S23E**, myasthenia gravis and multiple sclerosis share the most pathway genes relative to other conditions, but within drug networks, multiple sclerosis predictions use more similar genes to Alzheimer's disease than myasthenia gravis (**Figure S23D**). Taken together, these results suggested that druggable targets connected to distinct neurological diseases through shared pathway mechanisms, and provided further rationale for considering network-protein drug class effects in additional disease indications.

### Supplemental Tables and Figures

| Phase | Notes | Support |
|---|---|---|
| Preclinical | Complement activation has been established in multiple neurodegenerative diseases | [15–17] |
| Preclinical/ Clinical | Complement inhibitors approved for paroxysmal nocturnal hemoglobinuria (PNH) support the clinical utility of these molecules | [18–20] |
| Preclinical/ Clinical | Overall clinical development pipeline of complement inhibitors in myasthenia gravis | [21–24] |
| Clinical | Complement inhibitors and clinical development for cold agglutinin disease | [25–27] |
| Preclinical/ clinical | Complement inhibitors as therapeutically useful for macular degeneration | [28] |

**Table S4** – Preclinical support and clinical development for complement inhibitors

| Protein or group | Number of drug-phenotype associations | Unique phenotypes | Number of approved drugs | Number of approved drugs w/o the protein or group |
|---|---|---|---|---|
| Complement System | 1,945 | 17 | 312 | 551 |
| Chemokine system | 3,149 | 22 | 306 | 639 |
| NPY, NPY1R, NPY2R, NPY5R | 2,242 | 13 | 305 | 578 |
| CXCR3 | 443 | 2 | 305 | 75 |
| CXCR5 | 443 | 2 | 305 | 77 |
| CNR2 | 1,435 | 6 | 304 | 490 |

**Table S5. Novel drug classes and number of approved active ingredients for drug classes based on downstream proteins.** Complement system includes: C1QA,C1QB,C1QC,C1R,C1S,C3,C3AR1,C5,C5AR1
Chemokine system includes: CCR1,CCR2,CCR3,CCR4,CCR5,CCR6,CCR7,CCR8,CXCL1,CXCL10,CXCL11,CXCL12,CXCL13,CXCL16,CXCL2,CXCL3,CXCL5,CXCL6,CXCL8,CXCL9,CXCR1,CXCR2,CXCR3,CXCR4,CXCR5,CXCR6

| | Target | | Comparator | |
|---|---|---|---|---|
| **Study Name** | **Unique Brand Names** | **Unique Patients** | **Unique Brand Names** | **Unique Patients** |
| ALS – Complement System | 40 | 1312 | 38 | 2243 |
| ALS – CNR2 | 42 | 1374 | 32 | 2200 |
| ALS – NPY | 40 | 1106 | 41 | 2449 |
| ALS – Chemokine | 40 | 1115 | 45 | 2435 |
| ALS – CXCR3 | 49 | 2659 | 5 | 1127 |
| ALS – CXCR5 | 48 | 2646 | 6 | 1128 |
| PD –Complement System | 52 | 34457 | 59 | 36746 |
| MG – Complement System | 53 | 6027 | 61 | 11669 |
| MG – CXCR3 | 59 | 14477 | 7 | 6568 |
| MG – CXCR5 | 57 | 14399 | 9 | 6576 |

**Table S6** – **Number of brand names, patients for each study**

| **Study Name** | *Mean-Squared Error* | *Coefficient of determination* |
|---|---|---|
| ALS – Complement System | 0.32 | -0.37 |
| ALS – CNR2 | 0.33 | -0.39 |
| ALS – NPY | 0.29 | -0.35 |
| ALS – Chemokine | 0.30 | -0.35 |
| ALS – CXCR3 | 0.27 | -0.28 |
| ALS – CXCR5 | 0.26 | -0.26 |
| PD – Complement System | 0.38 | -0.50 |
| MG – Complement System | 0.31 | -0.35 |
| MG – CXCR3 | 0.31 | -0.43 |
| MG – CXCR5 | 0.30 | -0.39 |

**Table S7. Table Mean-Squared Error and Coefficient of Determination for all studies**

| | CS-class drugs with gene in top, over-expressed genes | Non-CS-class drugs with gene in top, over-expressed genes | Ratio in top | CS-class drugs with gene in bottom, under-expressed genes | Non-CS-class drugs with gene in bottom, under-expressed genes | Ratio in bottom |
|---|---|---|---|---|---|---|
| C5AR1 | 2 | 3 | 0.66 | 0 | 1 | - |
| C5 | 5 | 1 | 5 | 4 | 8 | 0.5 |
| C3AR1 | 4 | 1 | 4 | 2 | 3 | 0.66 |
| C3 | 5 | 4 | 1.25 | 6 | 8 | 0.75 |
| C1S | 7 | 4 | 1.75 | 1 | 3 | 0.33 |
| C1R | 11 | 8 | 1.375 | 2 | 5 | 0.4 |
| C1Q | 1 | 1 | 1 | 2 | 1 | 2 |
| C1QB | 2 | 1 | 2 | 2 | 4 | 0.5 |
| C1QA | 4 | 0 | - | 4 | 0 | - |

**Table S8. Complement system proteins in top or bottom gene expression for CS/non-CS class drugs from the PharmOmics dataset.** Ratio represents the number of "CS-class" drugs: number of "non-CS" drugs with the gene in the top ("ratio in top") or bottom ("ratio in bottom") of drug-induced gene changes.

| Domain | Notes | Support |
|---|---|---|
| Regulatory | FDA provded guidance and expcecttions about using RWD to generate RWE in translaitonal pipelines | [29,30] |
| Application | Accelerated approval as a single arm trial using a historical control | [31] |
| Application | Combining RWD with systems pharmacology to discern intrinsic factors for polatuzumab dosing | [32] |
| Application | Optimizing pediatric dosing for anemia patients | [33] |

**Table S9** – Impact of RWD on translational pipelines

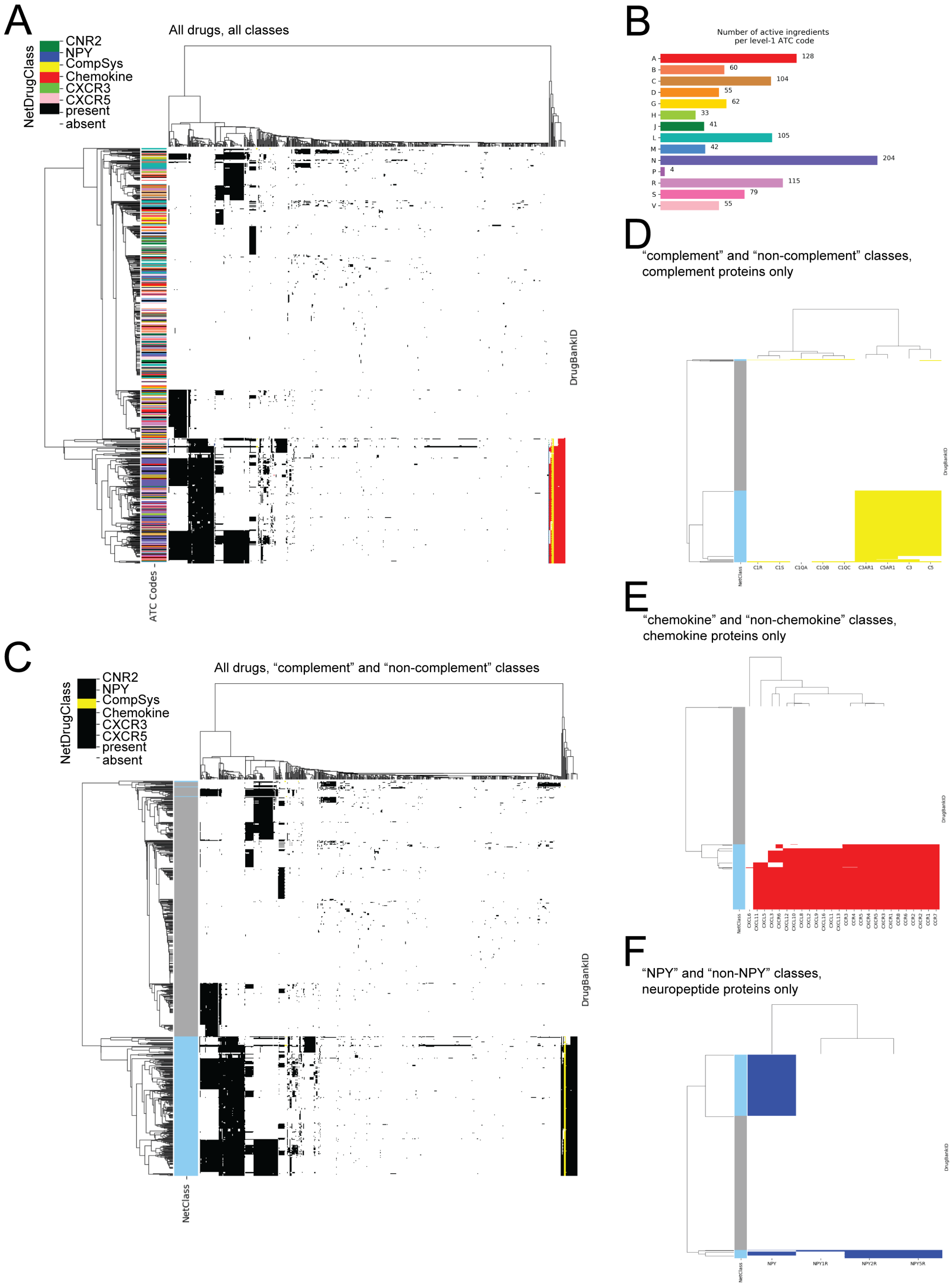
A
All drugs, all classes
NetDrugClass
CNR2
NPY
CompSys
Chemokine
CXCR3
CXCR5
present
absent
DrugBankID
ATC Codes
B
Number of active ingredients
per level-1 ATC code
A 128
B 60
C 104
D 55
G 62
H 33
J 41
L 105
M 42
N 204
P 4
R 115
S 79
V 55
C
All drugs, "complement" and "non-complement" classes
NetDrugClass
CNR2
NPY
CompSys
Chemokine
CXCR3
CXCR5
present
absent
DrugBankID
NetClass
D
"complement" and "non-complement" classes,
complement proteins only
DrugBankID
NetClass
C1R
C1S
C1QA
C1QB
C1QC
C3AR1
C5AR1
C3
C5
E
"chemokine" and "non-chemokine" classes,
chemokine proteins only
DrugBankID
NetClass
F
"NPY" and "non-NPY" classes,
neuropeptide proteins only
DrugBankID
NetClass
NPY
NPY1R
NPY2R
NPY5R

**Figure S1. ALS-PathFX finds broad associations of approved drugs to ALS pathways.** The heatmaps contains data for each drug (row) and network proteins (column) found in at least one approved drug association to an ALS gene pathway. In (**A**) row colors represent ATC codes and heatmap colors encode whether or not a protein is discovered in a drug network (black vs white), and additional colors represent if the network protein is part of a study class: CNR2 (dark green), neuropeptide Y (dark blue), complement system (yellow), chemokine (red), CXCR5 (light green), or CXCR3 (pink). All level-1 ATC codes and number of drugs associated with a code are depicted in (**B**). Additional heatmaps highlight drug network classes for all drugs and all of their network proteins considered in the complement system and non-complement system class (**C**), or highlight the class-specific proteins for the complement system class (**D**), chemokine class (**E**), and neuropeptide Y class (**F**).

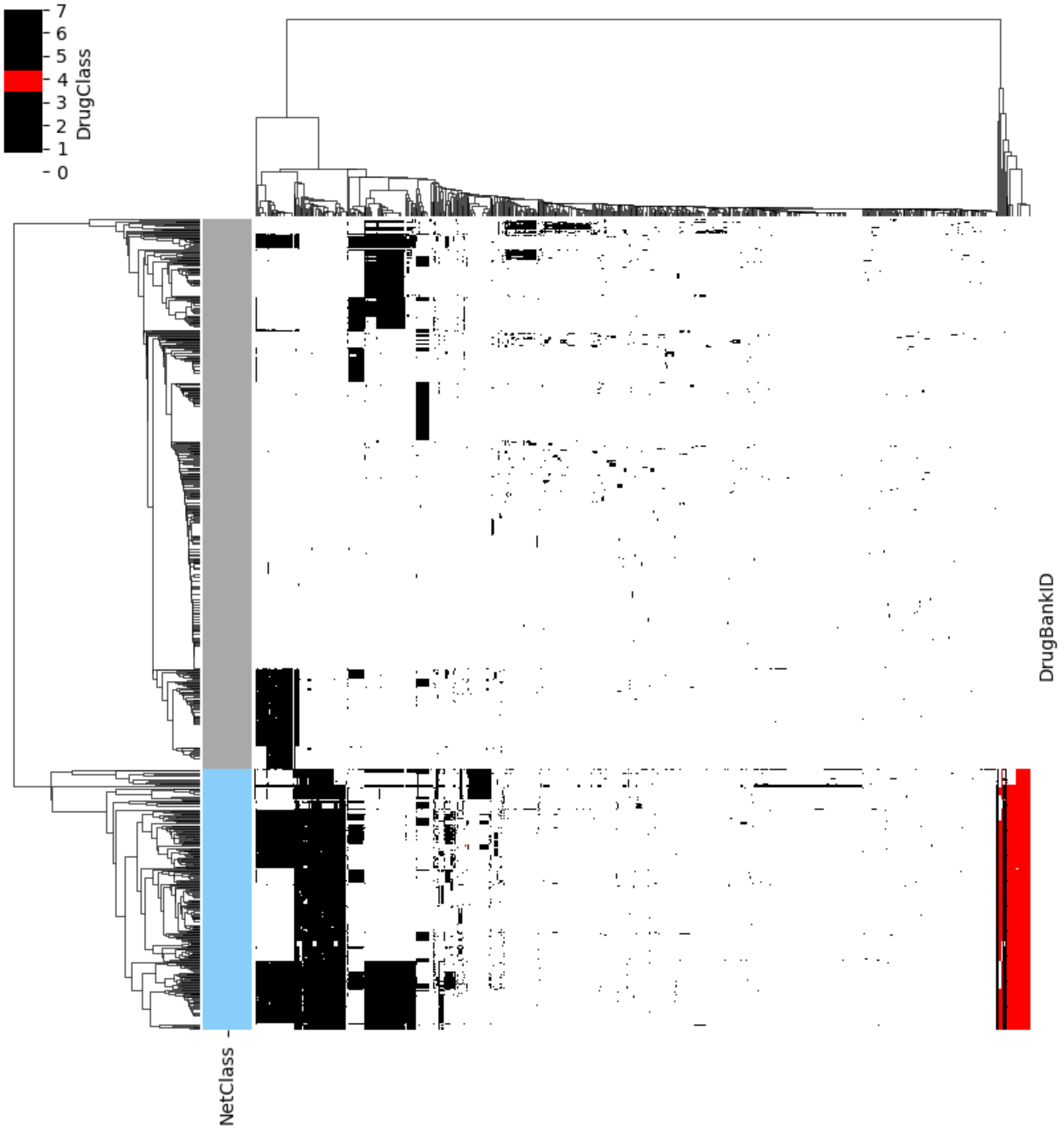

**Figure S2 – ALS-PathFX network proteins for drugs in the "chemokine" and "non-chemokine" drug classes**. The heatmap contains data for each drug (row) and network protein (column) found in at least one approved drug association to an ALS gene pathway. Heatmap colors encode whether or not a protein is discovered in a drug network (black vs white), and red indicates that the protein is in the chemokine network-drug class. Light blue/grey indicates whether the drug is in the target/comparator drug classes. Importantly, the non-chemokine class drugs contain no chemokine proteins (no red).

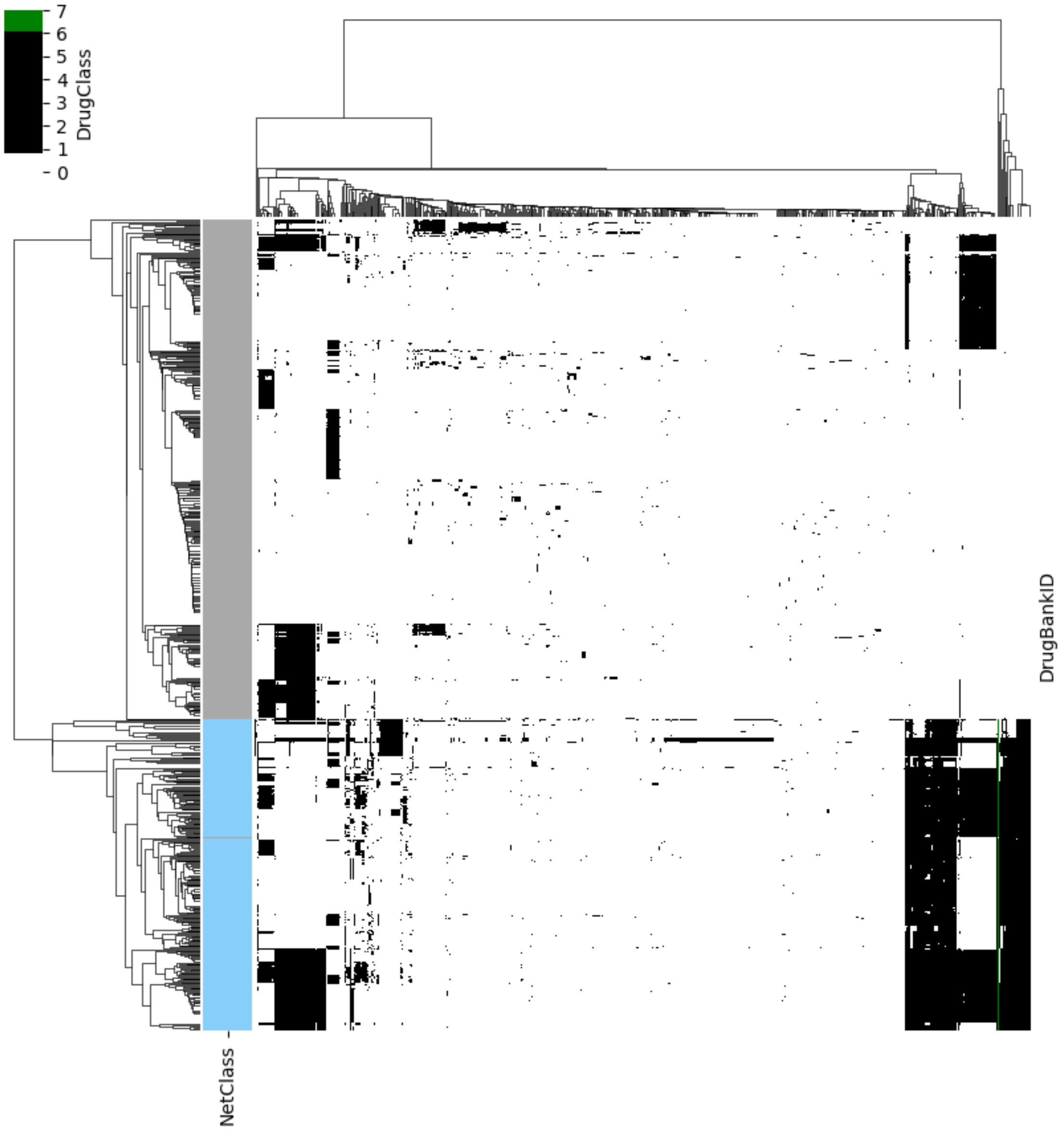

**Figure S3** – **ALS-PathFX network proteins for drugs in the "CNR2" and "non- CNR2" drug classes**. The heatmap contains data for each drug (row) and network protein (column) found in at least one approved drug association to an ALS gene pathway. Heatmap colors encode whether or not a protein is discovered in a drug network (black vs white), and green indicates that the protein is in the CNR2 network-drug class. Light blue/grey indicates whether the drug is in the target/comparator drug classes. Importantly, the non- CNR2 class drugs do not contain CNR2 (no green).

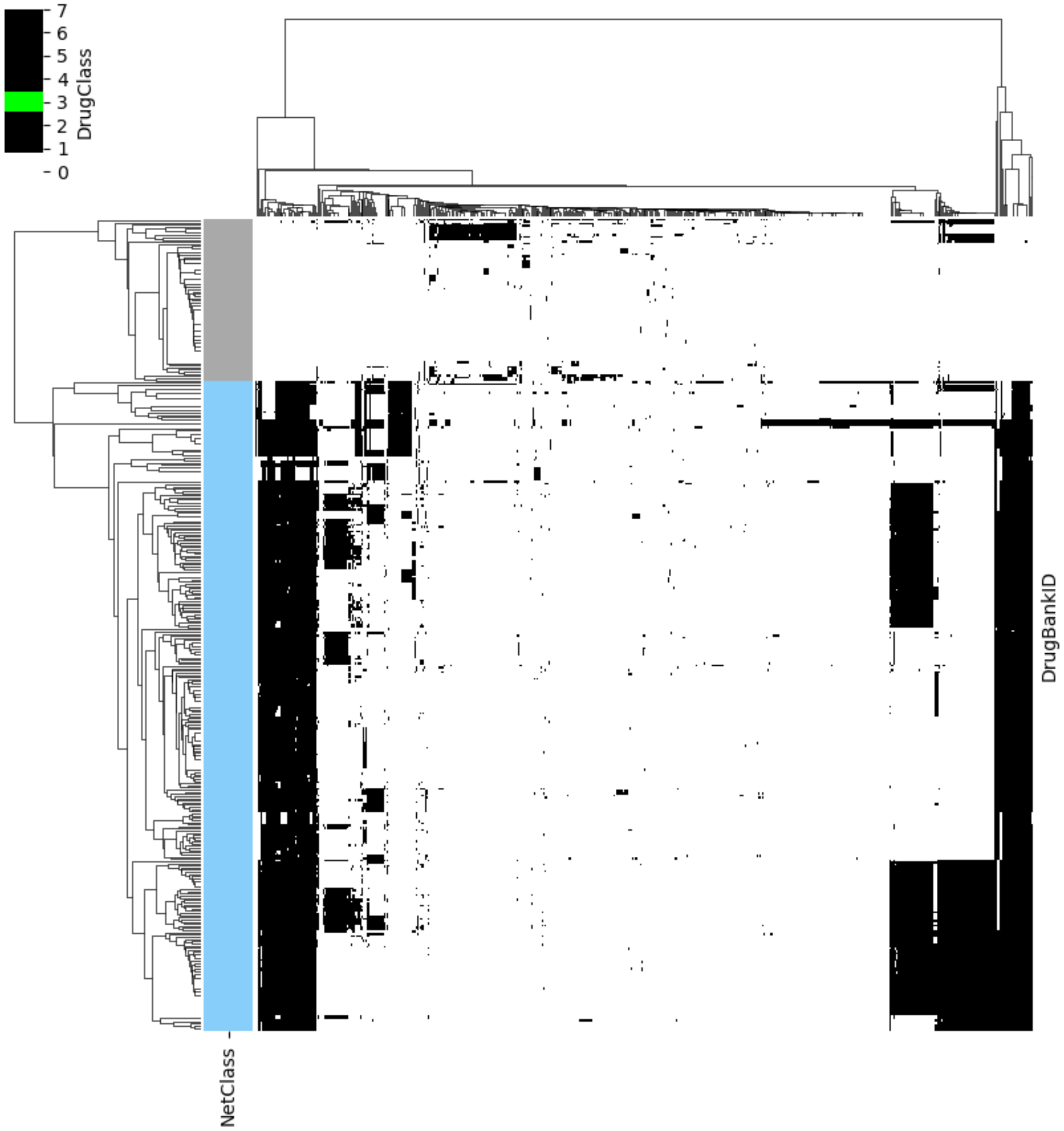

**Figure S4 – ALS-PathFX network proteins for drugs in the “CXCR3” and “non- CXCR3” drug classes**. The heatmap contains data for each drug (row) and network protein (column) found in at least one approved drug association to an ALS gene pathway. Heatmap colors encode whether or not a protein is discovered in a drug network (black vs white), and bright green indicates that the protein is in the CXCR3 network-drug class. Light blue/grey indicates whether the drug is in the target/comparator drug classes. Importantly, the non- CXCR3 class drugs do not contain CXCR3 (no bright green).

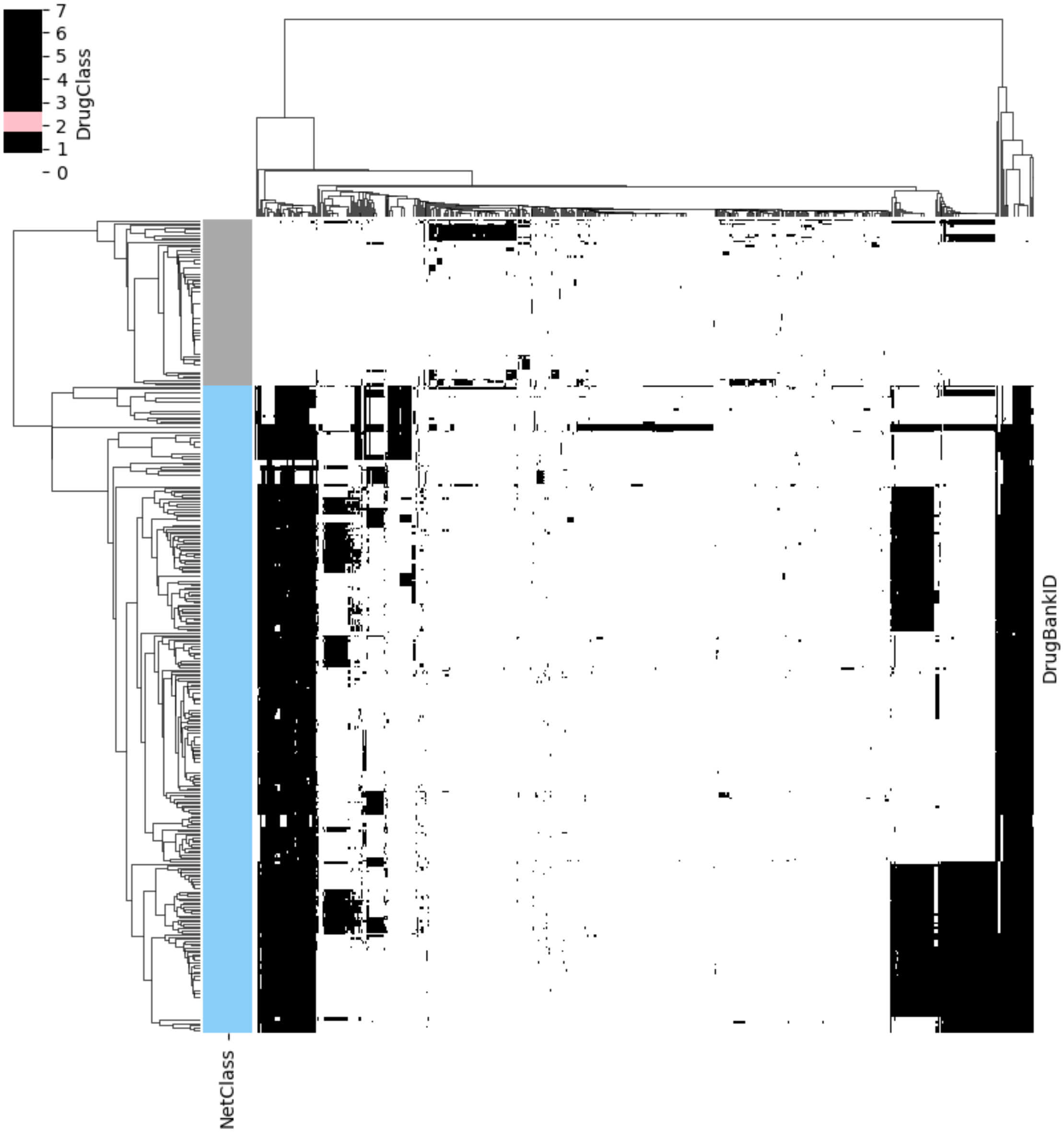

**Figure S5 – ALS-PathFX network proteins for drugs in the "CXCR5" and "non- CXCR5" drug classes**. The heatmap contains data for each drug (row) and network protein (column) found in at least one approved drug association to an ALS gene pathway. Heatmap colors encode whether or not a protein is discovered in a drug network (black vs white), and pink indicates that the protein is in the CXCR5 network-drug class. Light blue/grey indicates whether the drug is in the target/comparator drug classes. Importantly, the non- CXCR5 class drugs do not contain CXCR5 (no pink).

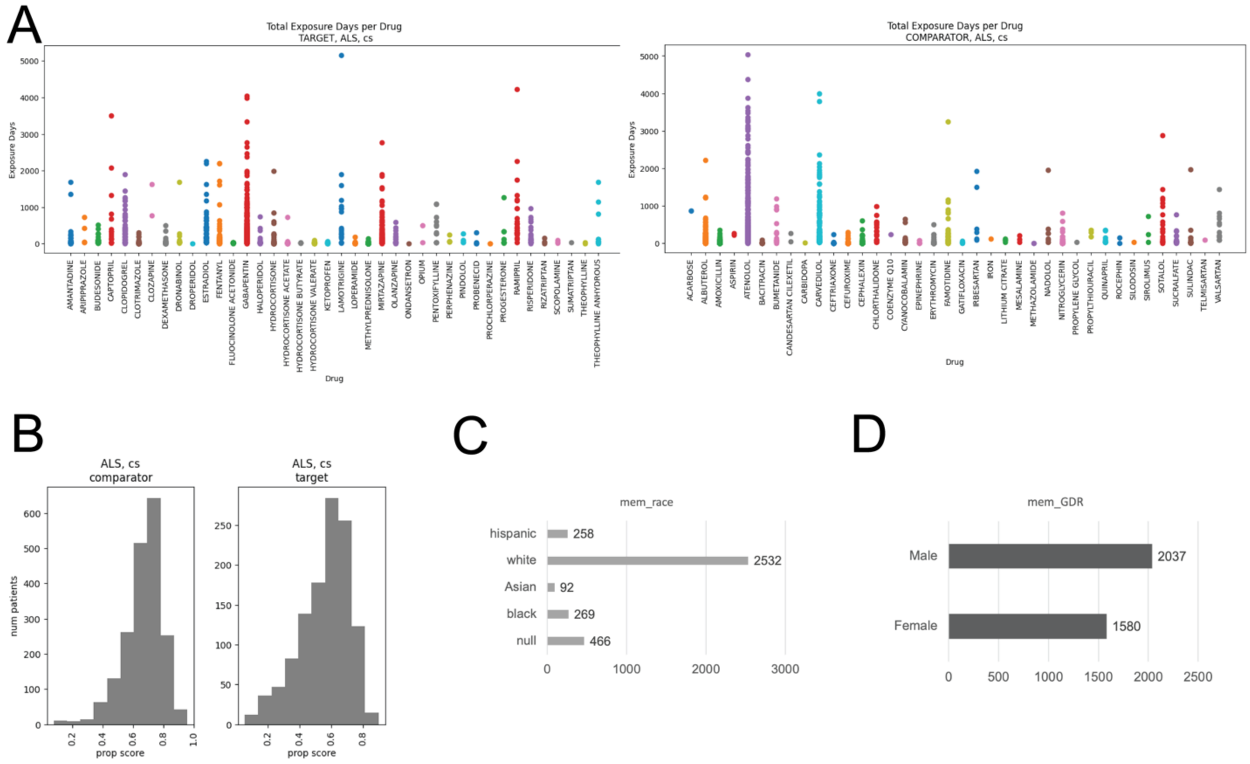

**Figure S6. Patient cohorts in network drug classes have sufficient covariate balance** (**A**) We plotted the total exposure days (sum of all prescriptions) per brand name per patient. We repeated this process for the target, CS-class (left) and comparator, non-CS-class (right) drugs. Each dot is a single patient. (**B**) The number of patients is plotted against their propensity score for patients on non-CS-class drugs (left) or CS-class drugs (right). The propensity score is the predicted probability using LogisticRegression on patient demographic, diagnostics, and medical prescription claims features. Patient demographics information for race (**C**) and gender (**D**) concepts.

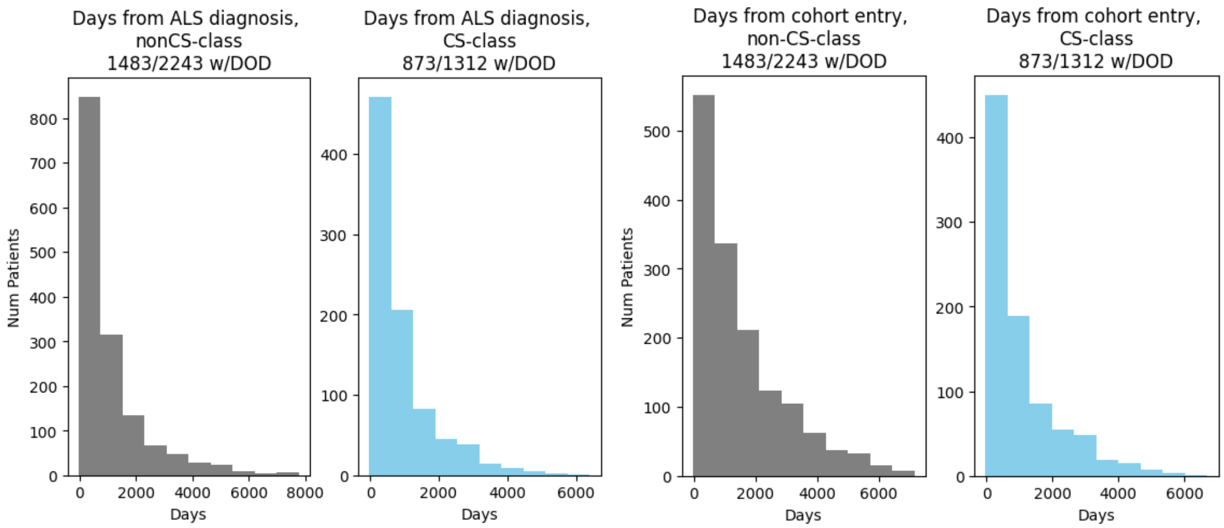

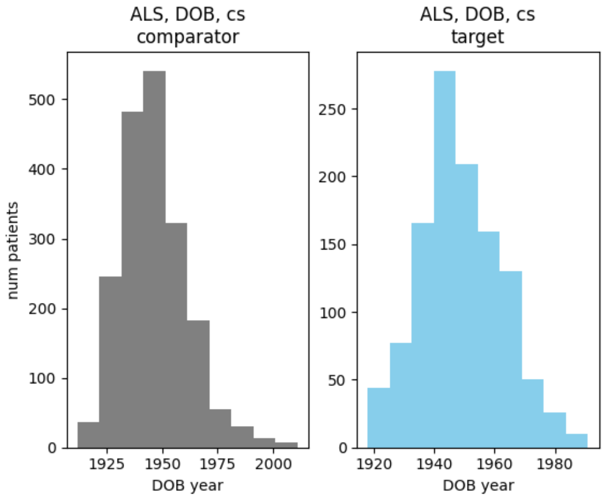

**Figure S7**. **Complement system target and comparator cohorts are not enriched for slow- or fast-responders, both have similar distributions of year-of-birth**. Time-to-death from first ALS diagnosis (top left) or from cohort entry (top right) in days is plotted for all patients on non-CS-class drugs (grey) and patients on CS-class drugs (skyblue). Cohort entry is defined as the time of exposure to first study drug (first RX claim). From first ALS diagnosis, the median and longest survival time for the non-CS-class and CS-class was 613 and 7,781 days (~21.3 years) and 552 and 6,390 days (~17.5 years), respectively (Kruskal stat = 8.79, p-value = 0.003). From cohort entry, the median and longest survival time for the non-CS-class and CS-class was 1,042 and 7,156 days (~19.6 years) and 617 and 6,691 days (~18.3 years), respectively (Kruskal stat = 78.22, p-value = $9.21 \times 10^{-19}$). There were 767 and 456 patients in the non-CS-class and CS-class who did not have a recorded date of death. The year-of-birth (DOB year) is plotted for the non-CS-class (grey) and CS-class (skyblue) cohorts (bottom center).

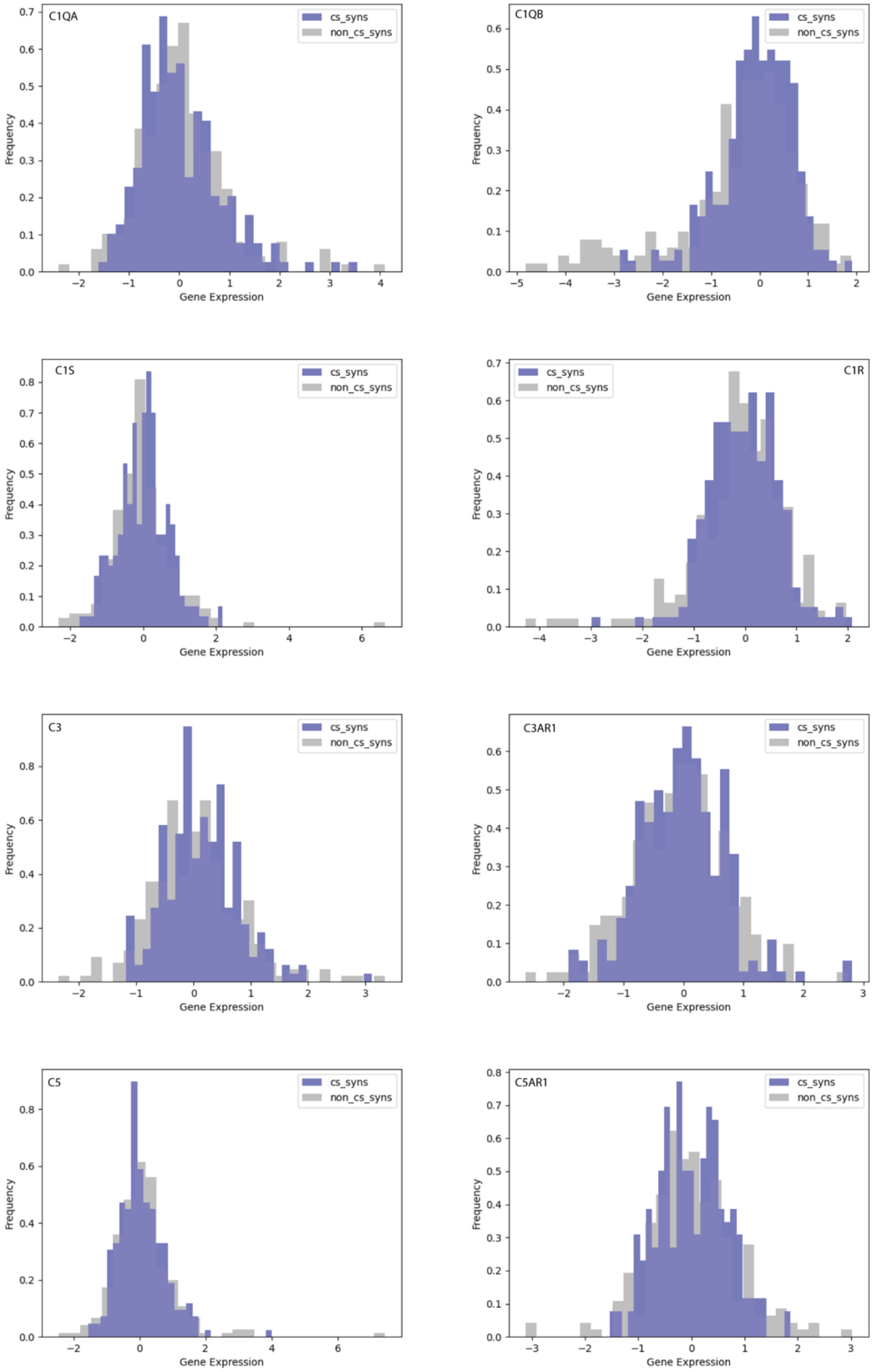

**Figure S8. LINCS level-5 gene expression shows no difference in complement system gene expression for either drug class.** The level-5, normalized gene expression score is plotted for 8 of the 9 complement system proteins where complement system ("cs_syns") and non-complement system ("non_cs_syns") drug classes are plotted in purple and grey, respectively.

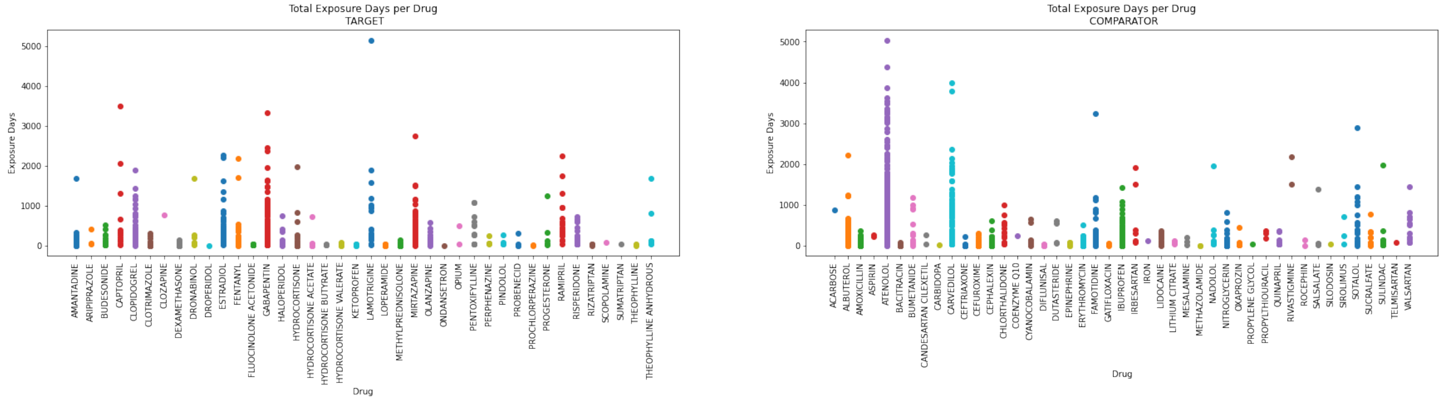

**Figure S9. Target/Comparator Drug Exposures for Chemokine Class in ALS**. We plotted the total exposure days (sum of all prescriptions) per brand name per patient. We repeated this process for the target (left) and comparator (right) drugs. Each dot is a single patient.

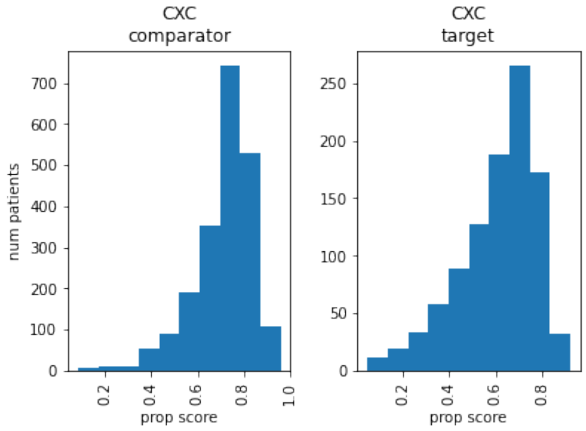

**Figure S10. Propensity score distributions for the Chemokine Class in ALS.** The number of patients is plotted against their propensity score for patients on non-network drug class drugs (left) or network-protein-class drugs (right). The propensity score is the predicted probability using LogisticRegression on patient demographic, diagnostics, and medical prescription claims features.

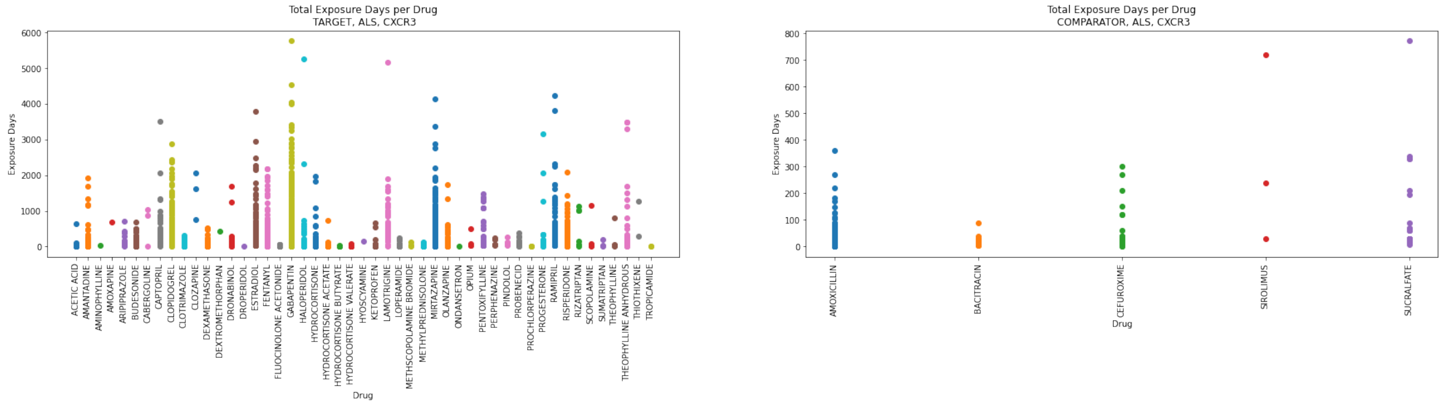

**Figure S11. Target/Comparator Drug Exposures for CXCR3 Class in ALS**. We plotted the total exposure days (sum of all prescriptions) per brand name per patient. We repeated this process for the target (left) and comparator (right) drugs. Each dot is a single patient.

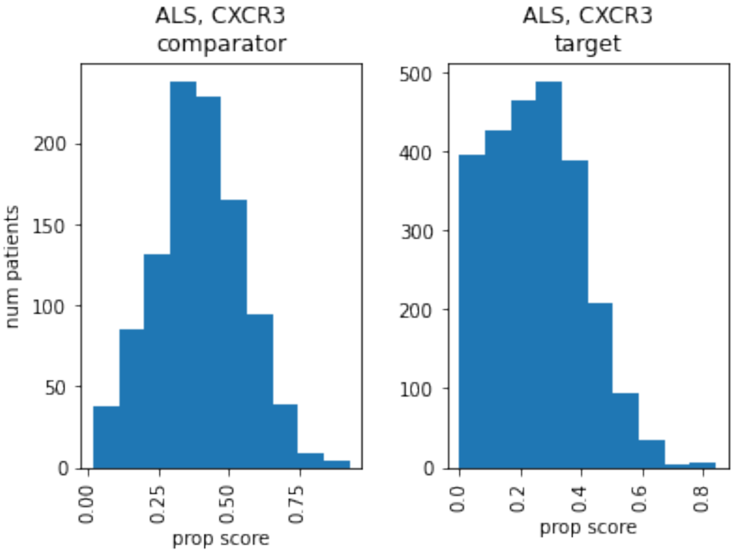

**Figure S12. Propensity score distributions for the CXCR3 Class in ALS.** The number of patients is plotted against their propensity score for patients on non-network drug class drugs (left) or network-protein-class drugs (right). The propensity score is the predicted probability using LogisticRegression on patient demographic, diagnostics, and medical prescription claims features.

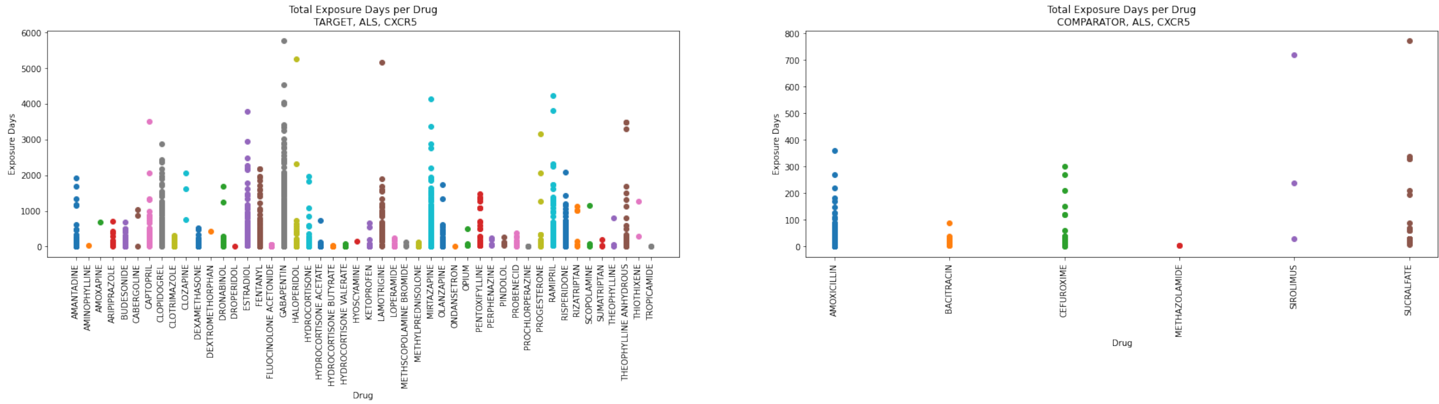

**Figure S13. Target/Comparator Drug Exposures for CXCR5 Class in ALS**. We plotted the total exposure days (sum of all prescriptions) per brand name per patient. We repeated this process for the target (left) and comparator (right) drugs. Each dot is a single patient.

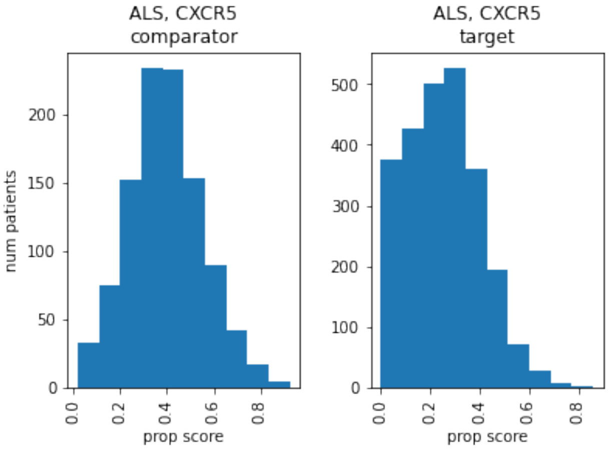

**Figure S14. Propensity score distributions for the CXCR5 Class in ALS.** The number of patients is plotted against their propensity score for patients on non-network drug class drugs (left) or network-protein-class drugs (right). The propensity score is the predicted probability using LogisticRegression on patient demographic, diagnostics, and medical prescription claims features.

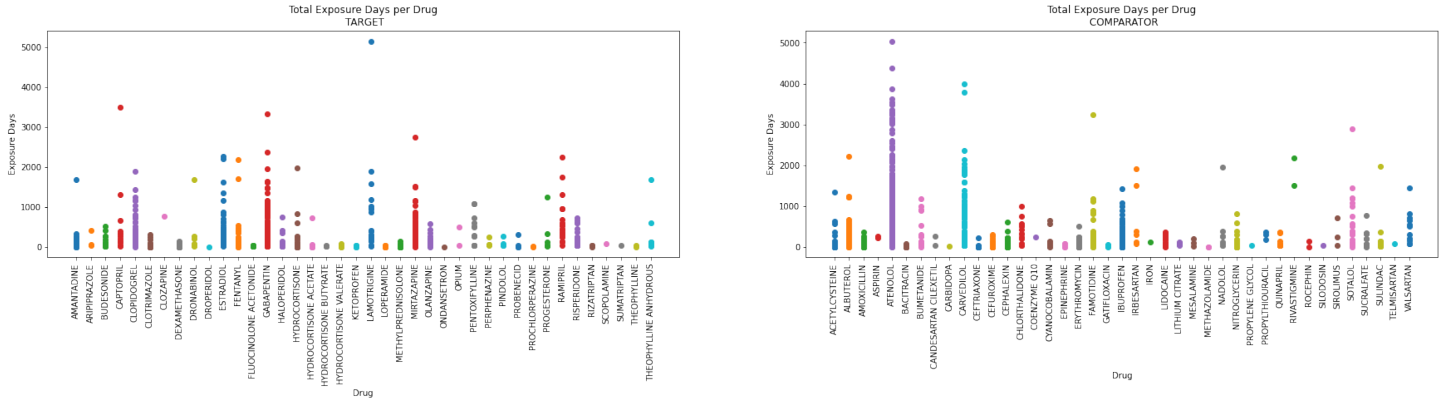

**Figure S15. Target/Comparator Drug Exposures for NPY Class in ALS**. We plotted the total exposure days (sum of all prescriptions) per brand name per patient. We repeated this process for the target (left) and comparator (right) drugs. Each dot is a single patient.

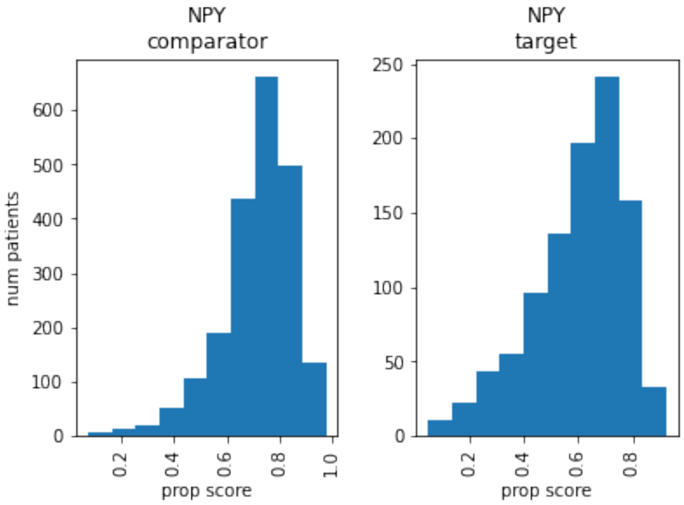

**Figure S16. Propensity score distributions for the NPY Class in ALS.** The number of patients is plotted against their propensity score for patients on non-network drug class drugs (left) or network-protein-class drugs (right). The propensity score is the predicted probability using LogisticRegression on patient demographic, diagnostics, and medical prescription claims features.

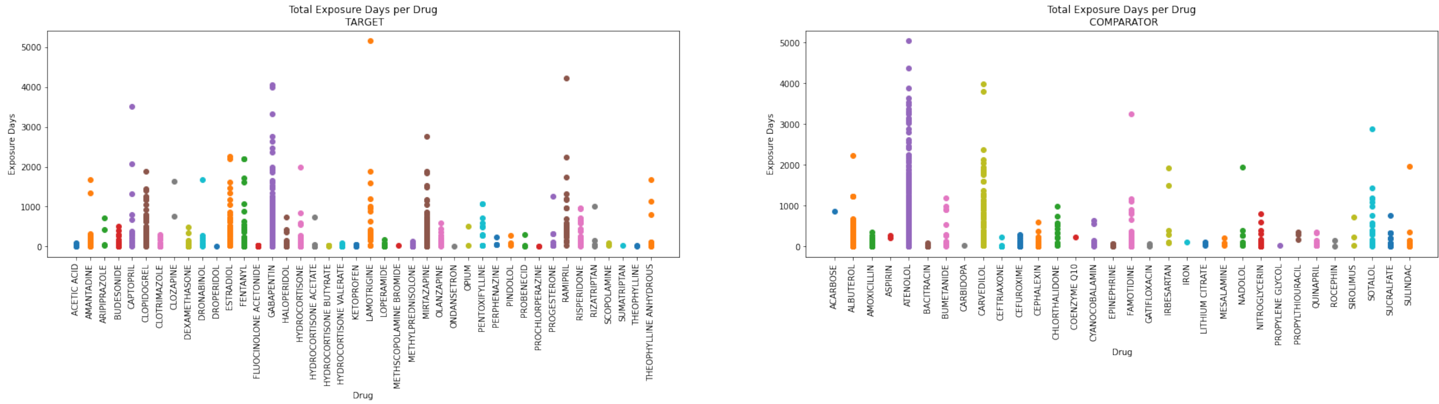

**Figure S17. Target/Comparator Drug Exposures for CNR2 Class in ALS**. We plotted the total exposure days (sum of all prescriptions) per brand name per patient. We repeated this process for the target (left) and comparator (right) drugs. Each dot is a single patient.

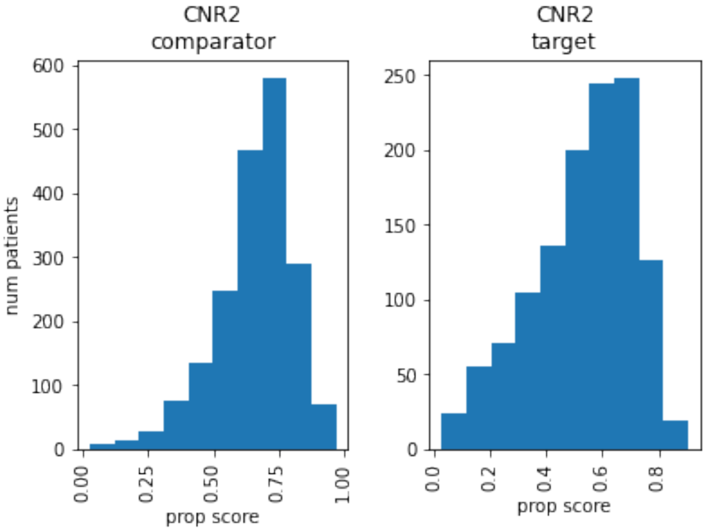

**Figure S18. Propensity score distributions for the CNR2 Class in ALS.** The number of patients is plotted against their propensity score for patients on non-network drug class drugs (left) or network-protein-class drugs (right). The propensity score is the predicted probability using LogisticRegression on patient demographic, diagnostics, and medical prescription claims features.

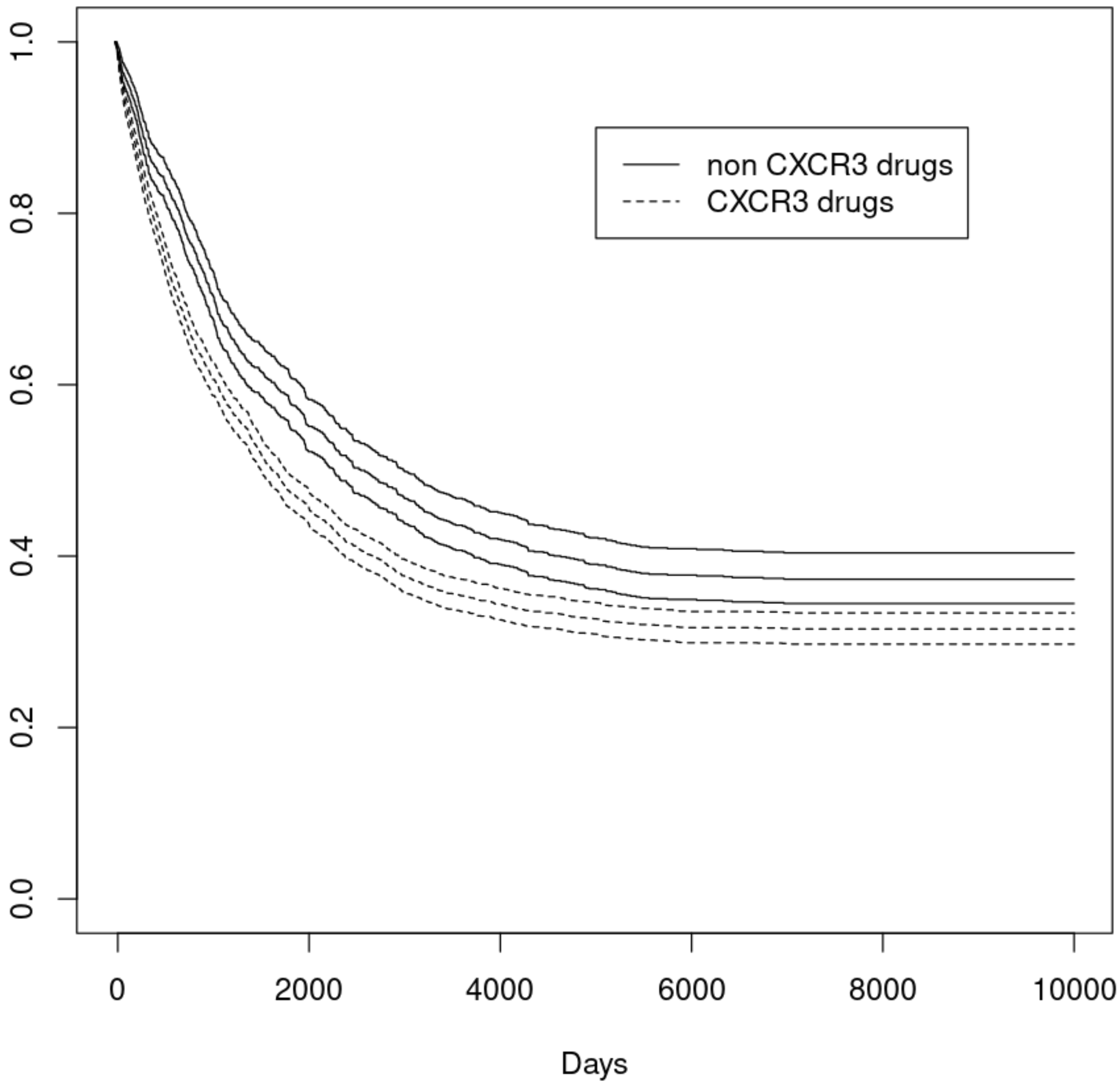

**Figure S19. Survival curve for CXCR3 class, ALS.** Proportion of the surviving population plotted against time in days. Patients exposed to a “CXCR3 class” or “NonCXCR3 class” drug are represented with a dotted or solid line, respectively. Survival curve includes 95% confidence interval.

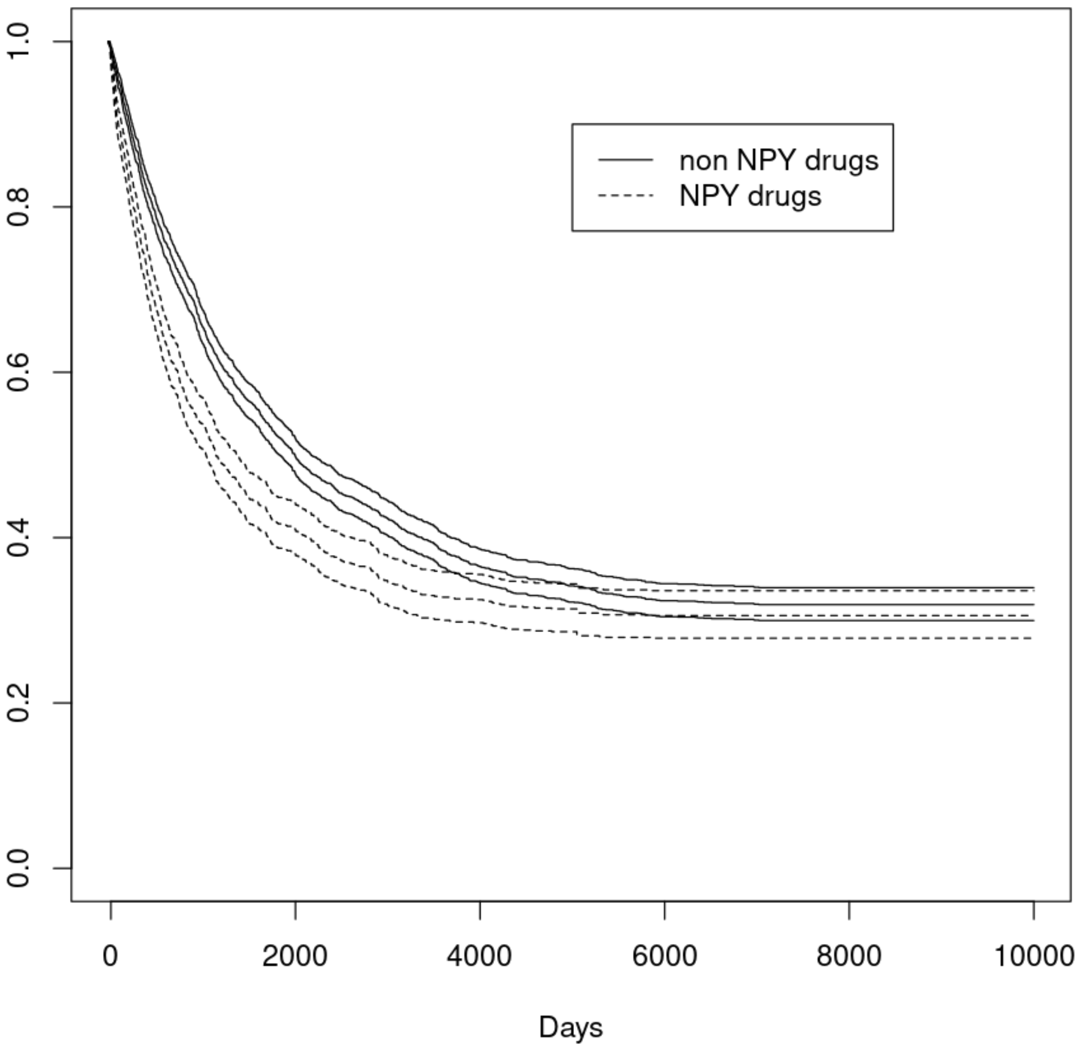

**Figure S20. Survival curve for NPY class, ALS.** Proportion of the surviving population plotted against time in days. Patients exposed to a “NPY class” or “NonNPY class” drug are represented with a dotted or solid line, respectively. Survival curve includes 95% confidence interval.

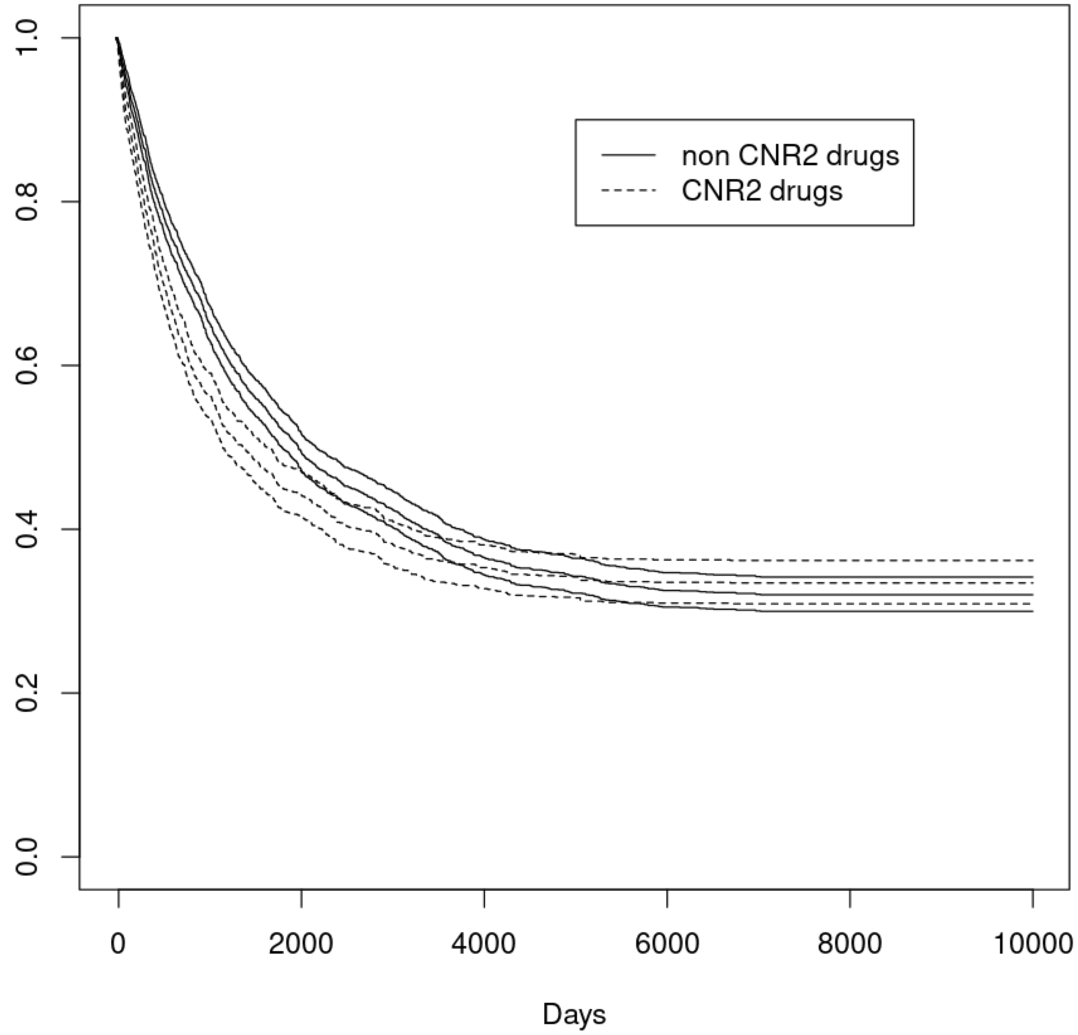

**Figure S21. Survival curve for CNR2 class, ALS.** Proportion of the surviving population plotted against time in days. Patients exposed to a “CNR2 class” or “NonCNR2 class” drug are represented with a dotted or solid line, respectively. Survival curve includes 95% confidence interval.

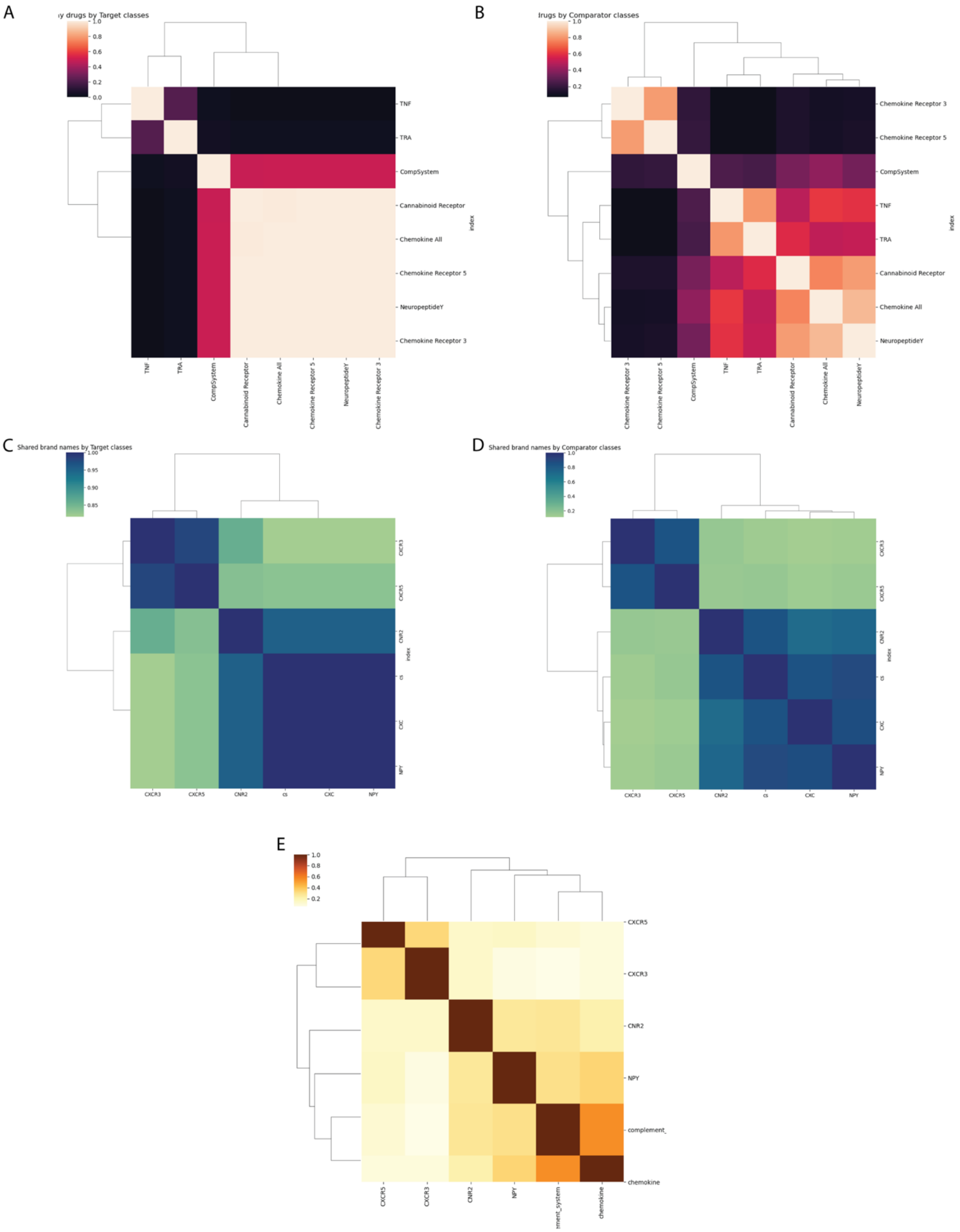
A
B
C
D
E
Shared brand names by Target classes
Shared brand names by Comparator classes
TNF
TRA
CompSystem
Cannabinoid Receptor
Chemokine All
Chemokine Receptor 5
NeuropeptideY
Chemokine Receptor 3
index
CXCR3
CXCR5
CNR2
cs
CXC
NPY
complement_
chemokine

**Figure S22.** Network classes have shared and distinct drugs, brand names, and disease pathway associations. Jaccard similarity of approved drugs in a network (**A**) or non-network class (**B**) and brand names discovered in ALS patient claims for the target (**C**) and comparator (**D**) classes. Jaccard similarity of disease pathways associated with each network class (**E**). CXCR3 = chemokine receptor 3, CXCR5 = chemokine receptor 5, cs = complement system, CNR2 = cannabinoid receptor 2, CXC = chemokine class, NPY = neuropeptide Y.

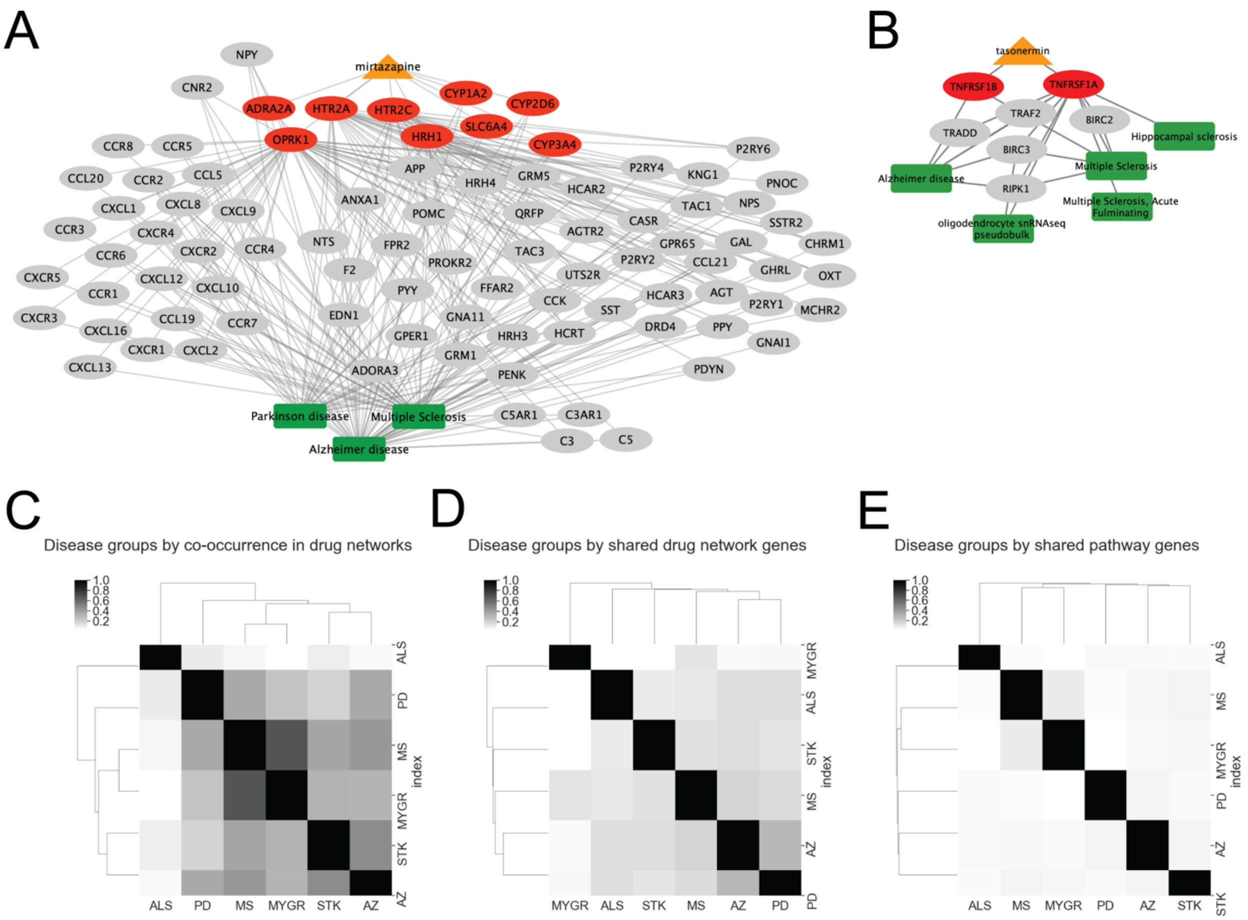

**Figure S23. Neurodegenerative pathway genes are shared within networks for approved drugs.** We provided two example network images where neurodegenerative pathway phenotypes are predicted in the same drug network: mirtazapine (**A**) and tasonermin (**B**). In both images, drugs, drug target proteins, downstream proteins, and pathway phenotypes are represented by orange triangles, red ellipses, grey ellipses, and green boxes, respectively. We quantified how often the neurodegenerative diseases co-occurred in drug networks using Jaccard similarity (number of drugs in which the diseases co-occur / number of drug networks with either disease) (**C**). We also quantified the network protein similarity (intersection of in-drug-network proteins / union of in-drug-network proteins) (**D**) and shared pathway genes (intersection of all disease genes / union of all disease genes) (**E**). Pathway genes are all genes associated with phenotypes in PathFX, but not all are used in a given network prediction. Diseases were visualized at the group level where ALS = Amyotrophic lateral sclerosis, PD = Parkinson's Disease, MS = Multiple Sclerosis, MYGR = Myasthenia Gravis, STK = Stroke, AZ = Alzheimer's Disease. All disease phenotypes belonging to a group are included in **Table S7.**

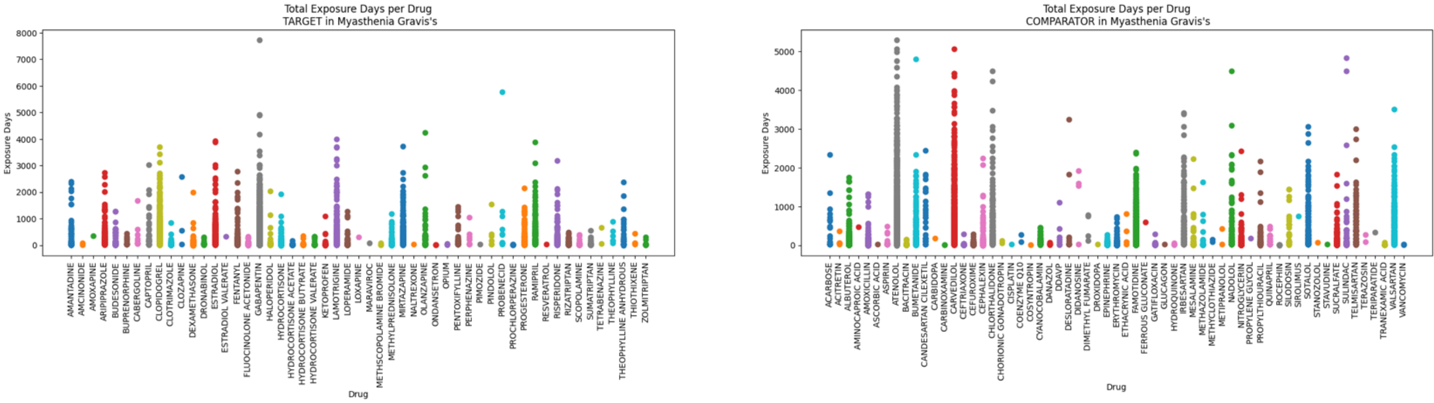

**Figure S24. Target/Comparator Drug Exposures for CS Class in Myasthenia Gravis**. We plotted the total exposure days (sum of all prescriptions) per brand name per patient. We repeated this process for the target (left) and comparator (right) drugs. Each dot is a single patient.

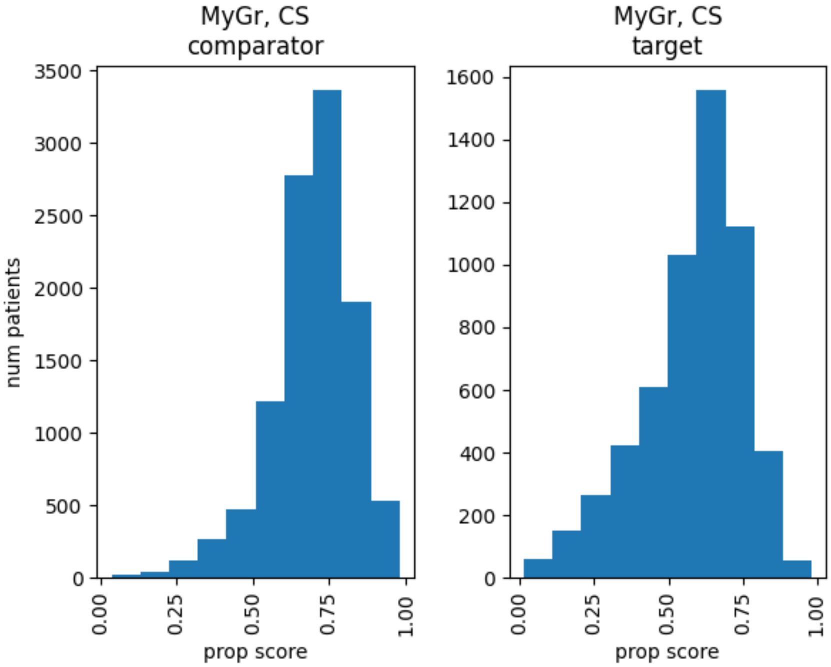

**Figure S25. Propensity score distributions for the CS Class in Myasthenia Gravis.** The number of patients is plotted against their propensity score for patients on non-network drug class drugs (left) or network-protein-class drugs (right). The propensity score is the predicted probability using LogisticRegression on patient demographic, diagnostics, and medical prescription claims features.

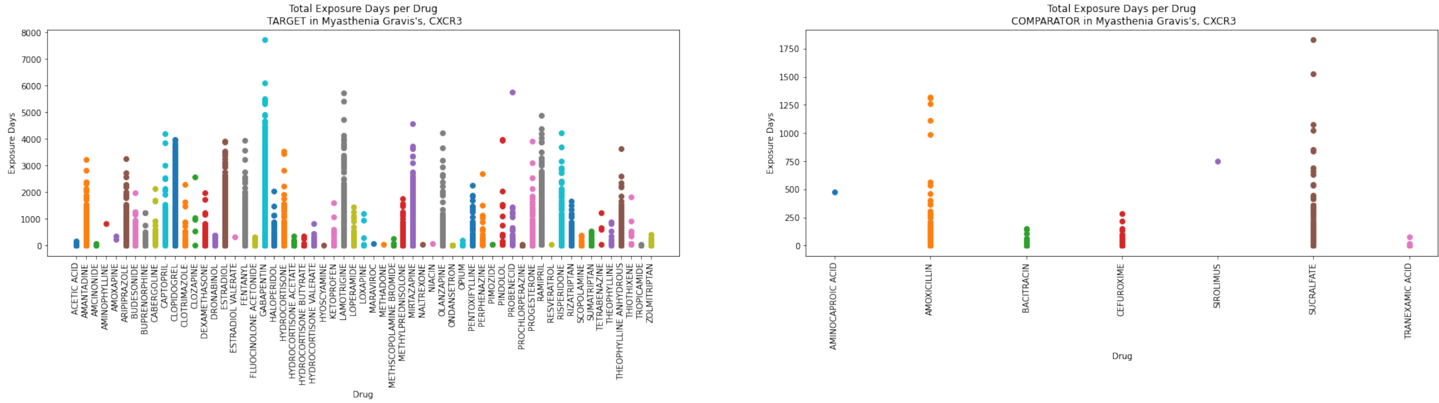

**Figure S26. Target/Comparator Drug Exposures for CXCR3 Class in Myasthenia Gravis**. We plotted the total exposure days (sum of all prescriptions) per brand name per patient. We repeated this process for the target (left) and comparator (right) drugs. Each dot is a single patient.

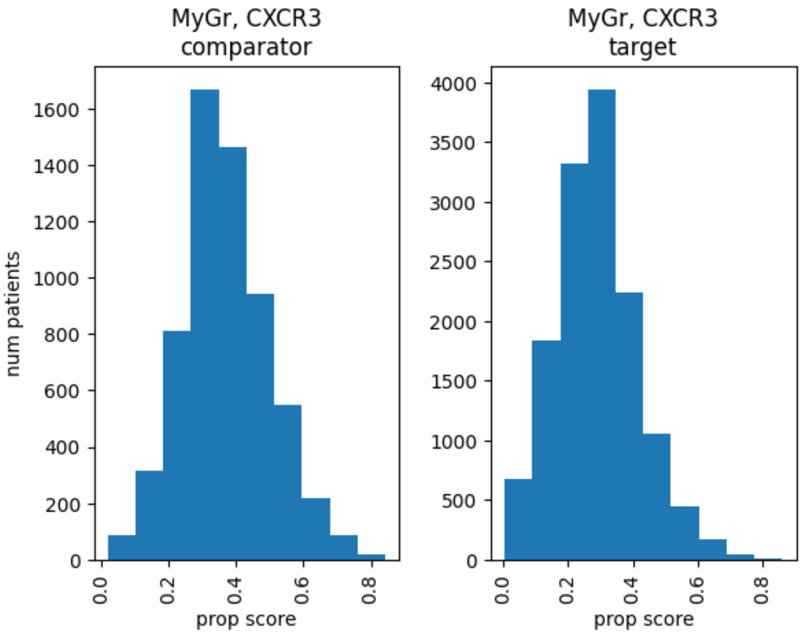

**Figure S27. Propensity score distributions for CXCR3 Class in Myasthenia Gravis.** The number of patients is plotted against their propensity score for patients on non-network drug class drugs (left) or network-protein-class drugs (right). The propensity score is the predicted probability using LogisticRegression on patient demographic, diagnostics, and medical prescription claims features.

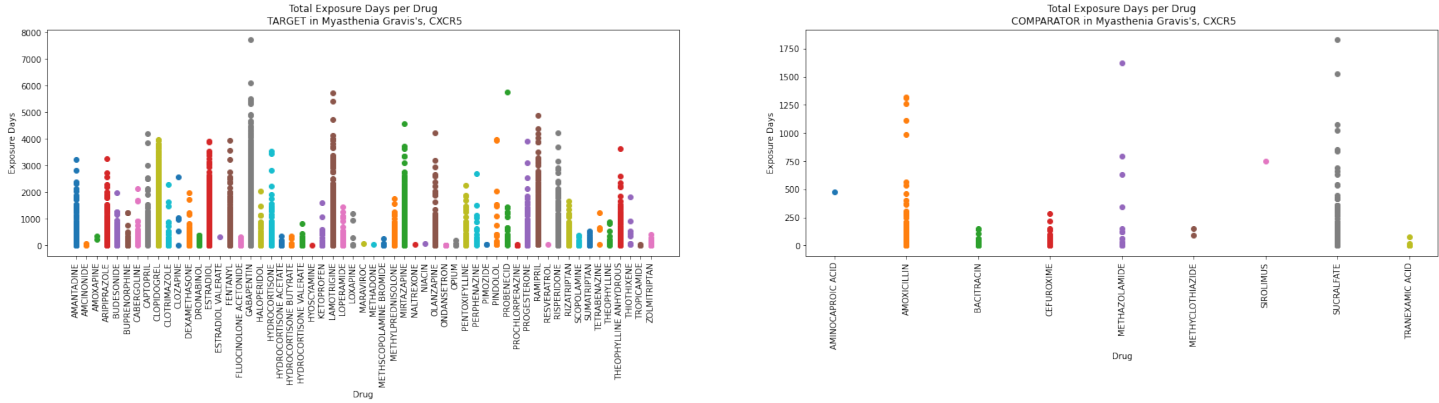

**Figure S28. Target/Comparator Drug Exposures for CXCR5 Class in Myasthenia Gravis**. We plotted the total exposure days (sum of all prescriptions) per brand name per patient. We repeated this process for the target (left) and comparator (right) drugs. Each dot is a single patient.

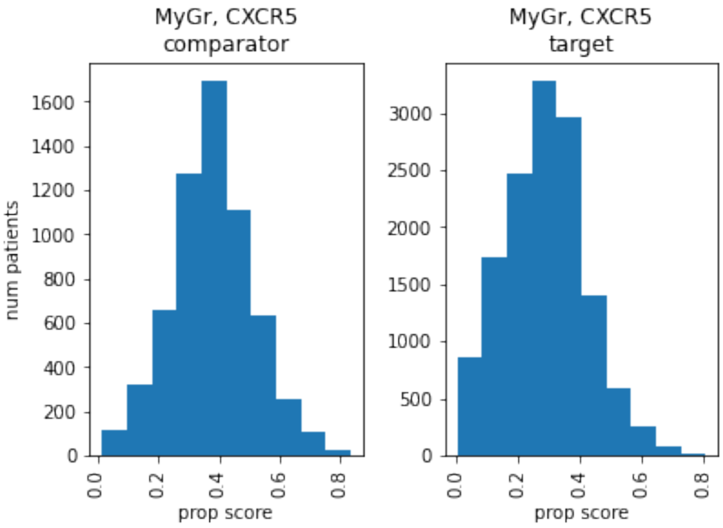

**Figure S29. Propensity score distributions for the CXCR5 Class in Myasthenia Gravis.** The number of patients is plotted against their propensity score for patients on non-network drug class drugs (left) or network-protein-class drugs (right). The propensity score is the predicted probability using LogisticRegression on patient demographic, diagnostics, and medical prescription claims features.

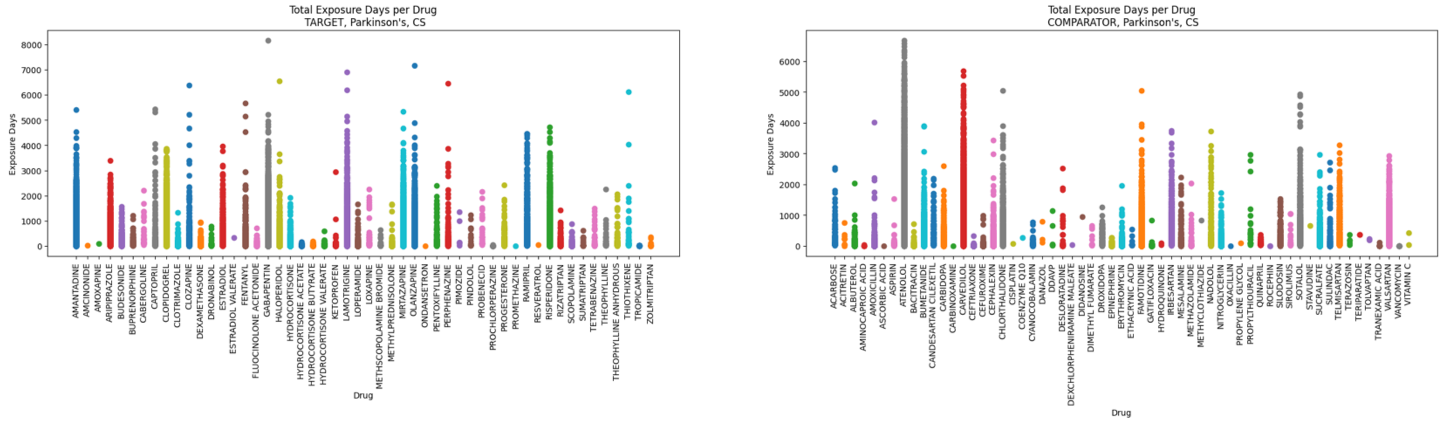

**Figure S30. Target/Comparator Drug Exposures for CS Class in Parkinson's disease**. We plotted the total exposure days (sum of all prescriptions) per brand name per patient. We repeated this process for the target (left) and comparator (right) drugs. Each dot is a single patient.

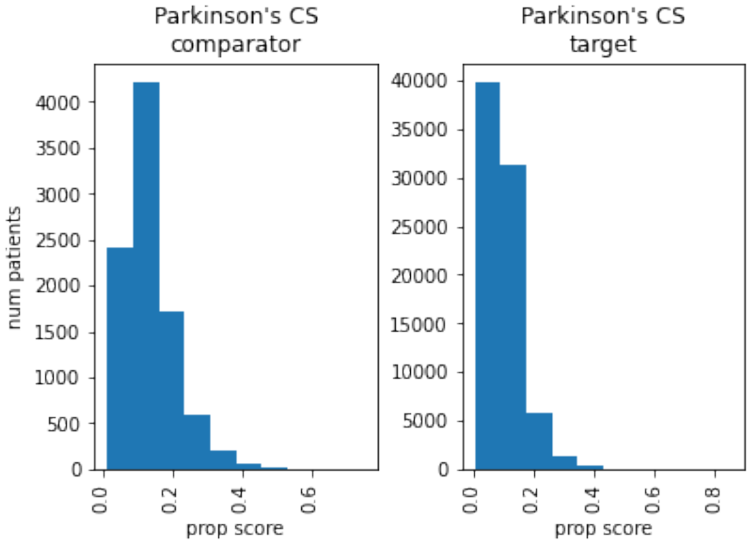

**Figure S31. Propensity score distributions for CS Class in Parkinson's disease.** The number of patients is plotted against their propensity score for patients on non-network drug class drugs (left) or network-protein-class drugs (right). The propensity score is the predicted probability using LogisticRegression on patient demographic, diagnostics, and medical prescription claims features.

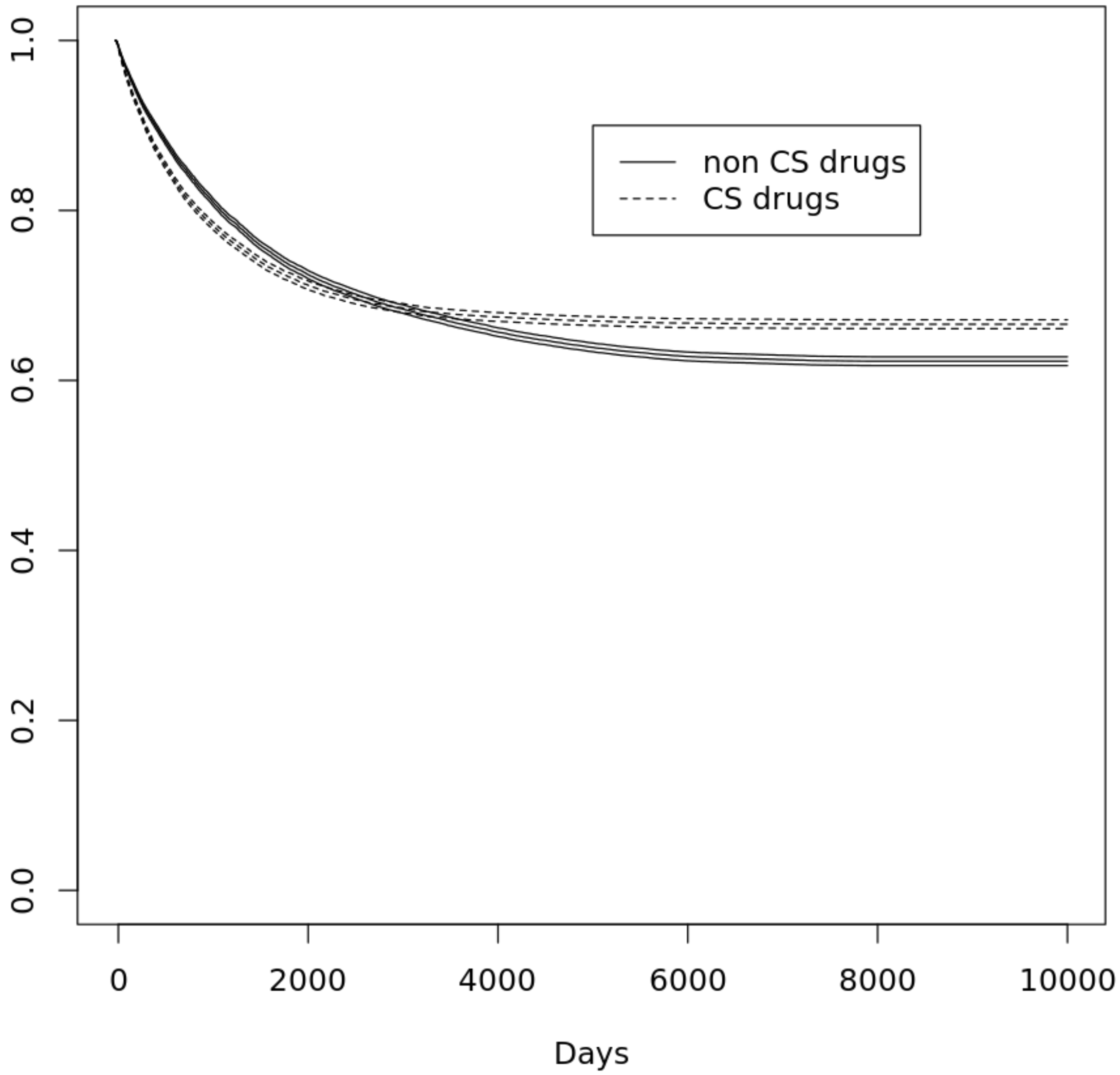

**Figure S32. Survival curve for CS class, Parkinson's.** Proportion of the surviving population plotted against time in days. Patients exposed to a "CS class" or "NonCS class" drug are represented with a dotted or solid line, respectively. Survival curve includes 95% confidence interval.
